# The impact of major depressive disorder on glycaemic control in type 2 diabetes: a longitudinal cohort study using UK Biobank primary care records

**DOI:** 10.1101/2022.01.13.22269229

**Authors:** Alexandra C Gillett, Saskia P Hagenaars, Dale Handley, Francesco Casanova, Katherine G Young, Harry Green, Cathryn M Lewis, Jess Tyrrell

## Abstract

**Background:** This study evaluates longitudinal associations between glycaemic control (mean and within-patient variability of glycated haemaglobin (HbA1c) levels) in individuals with type 2 diabetes (T2D) and major depressive disorder (MDD), focusing on the timings of these diagnoses.

**Methods:** In UK Biobank, T2D was defined using self-report and linked health outcome data, then validated using polygenic scores. Repeated HbA1c measurements (mmol/mol) over the 10 years following T2D diagnosis were outcomes in mixed effects models, with T2D disease duration included using restricted cubic splines. Four MDD exposures were considered: MDD diagnosis prior to T2D diagnosis (pre-T2D MDD), time between pre-T2D MDD diagnosis and T2D, new MDD diagnosis during follow-up (post-T2D MDD) and time since post-T2D MDD diagnosis. Models with and without covariate adjustment were considered.

**Results:** T2D diagnostic criteria were robustly associated with T2D polygenic scores. In 11,837 T2D cases (6.9 year median follow-up), pre-T2D MDD was associated with a 0.92 increase in HbA1c (95% CI: [0.00, 1.84]), but earlier pre-T2D MDD diagnosis correlated with lower HbA1c. These pre-T2D MDD effects became non-significant after covariate adjustment. Post-T2D MDD individuals demonstrated increasing HbA1c with years since MDD diagnosis (/3 = 0.51, 95% CI: [0.17, 0.86]). Retrospectively, looking across all follow-up, within-patient variability in HbA1c was 1.16 (95% CI: 1.13-1.19) times higher in post-T2D MDD.

**Conclusions:** The timing of MDD diagnosis is important for understanding glycaemic control in T2D. Poorer control was observed in MDD diagnosed post-T2D, highlighting the importance of depression screening in T2D, and closer monitoring for individuals who develop MDD after T2D.

## Background

Major depressive disorder (MDD) and type 2 diabetes (T2D) are substantial global health burdens, occurring together at twice the frequency expected by chance^1^. Adults with MDD have a 37% higher risk of developing T2D^2^, and people with T2D face a 15% higher risk of developing MDD^3^.

For individuals with T2D, comorbid depression is associated with elevated risk of diabetic complications^4^ and all-cause mortality^5^. The underlying mechanisms between these disorders and outcomes remain poorly understood, but could include lifestyle factors, non-adherence to T2D treatment, use of antidepressant medication or genetic factors^1^.

One potential link between MDD and adverse outcomes in T2D is glycaemic control, with glycated haemoglobin (HbA1c) representing a reliable measure of long-term glycemia. Elevated and more variable HbA1c levels are associated with increased risk of long-term diabetic complications, such as stroke and cardiovascular disease^6–8^. Prior studies have shown mixed support for the association between MDD and HbA1c^1,9^, but many are limited to a cross-sectional design. A meta-analysis that considered longitudinal effects of depressive symptoms on HbA1c found that depressive symptoms associated with higher HbA1c, and so poorer glycaemic control, across a mean follow-up of 3 years (*n*=3683 across six studies)^9^.

Furthermore, the relative timing of MDD and T2D diagnoses may also impact T2D clinical characteristics. A cross-sectional study showed that depression diagnosed after T2D is linked with poorer glycaemic control and a higher prevalence of diabetic complications, compared to T2D patients with no or with pre-existing depression^10^.

Understanding the impact of MDD on glycaemic control across time is crucial for delivering appropriate clinical care to individuals living with both MDD and T2D. Longitudinal studies to date have not explored the relationship between the relative diagnostic timings of these disorders and HbA1c trends over time. To address this gap, our study uses UK Biobank (UKB) primary care records to perform extensive longitudinal modelling of HbA1c in people with T2D in a retrospective, observational study, using information on MDD diagnosis. In the UK, HbA1c is measured by a general practitioner (GP) every 3-6 months in individuals with T2D^11^. Therefore, the linked primary care data available in UKB provides a unique opportunity to test the longitudinal relationship between HbA1c (mean levels and variability) and MDD over a 10-year period following T2D diagnosis. Our analysis incorporates four MDD exposures:

1. pre-T2D MDD diagnosis (ever diagnosed with MDD prior to T2D),
2. time between pre-T2D MDD diagnosis and T2D,
3. time-varying post-T2D MDD (newly diagnosed with MDD during follow-up period), and
4. time since post-T2D MDD diagnosis.

By considering these exposures, we aim to gain a deeper understanding of how MDD and its timing in relation to T2D affect glycaemic control.

## Methods

### Study population

The UK Biobank (UKB) is a health study of ∼500,000 individuals recruited between 2006 and 2010 in the United Kingdom, aged 40-70^12^. Linked primary care records are available for ∼230,000 individuals (46%), encompassing clinical events, blood test results and prescriptions, providing longitudinal patient information^13^.

#### T2D Classification and Validation

UKB participants with primary care records were classified into T2D cases and controls. T2D cases met specific T2D diagnostic criteria (detailed below), while controls did not. The T2D diagnostic criteria were validated using T2D polygenic scores (PGSs) in European ancestry participants meeting genetic quality control criteria^14^ (Supplementary Methods (SuppMethods) 1).

#### Type 2 diabetes (T2D) diagnostic criteria

T2D cases were identified based on the presence of any two of the following: 1) a primary care diagnosis code for T2D (Supplementary Table (SuppTable) S1), 2) an ICD9/ICD10 diagnosis code for T2D (SuppTable S2), 3) any HbA1c measurement > 48 mmol/mol (6.5%), 4) any prescription for glucose lowering medication (SuppMethods 2), and 5) a self-reported diagnosis for T2D with reported age at onset > 35 years. T2D diagnosis date was then the earliest occurrence (Figure 2). T2D cases were excluded if they had a primary care code specific to type 1 diabetes, an insulin prescription within a year of T2D diagnosis, or a prescription for multiple diabetic medications at T2D diagnosis (SuppMethods 2-3). Individuals prescribed one diabetic medication (monotherapy) at T2D diagnosis were not excluded.

#### Exclusion criteria for longitudinal analysis

For the longitudinal analysis, we excluded participants meeting any of the following criteria: 1) were a T2D control, 2) had fewer than two HbA1c measurements after T2D diagnosis, 3) had age at T2D diagnosis < 18 years, 4) had no recorded HbA1c measurements > 38 mmol/mol (3.5%)^15^ within a six-month window of T2D diagnosis, 5) had diagnostic codes for bipolar, psychotic or substance-use disorders^16^, 6) had a MDD diagnosis without a specified diagnosis date, or 7) were missing self-reported ethnicity (Figure 1).

**Figure 1.**
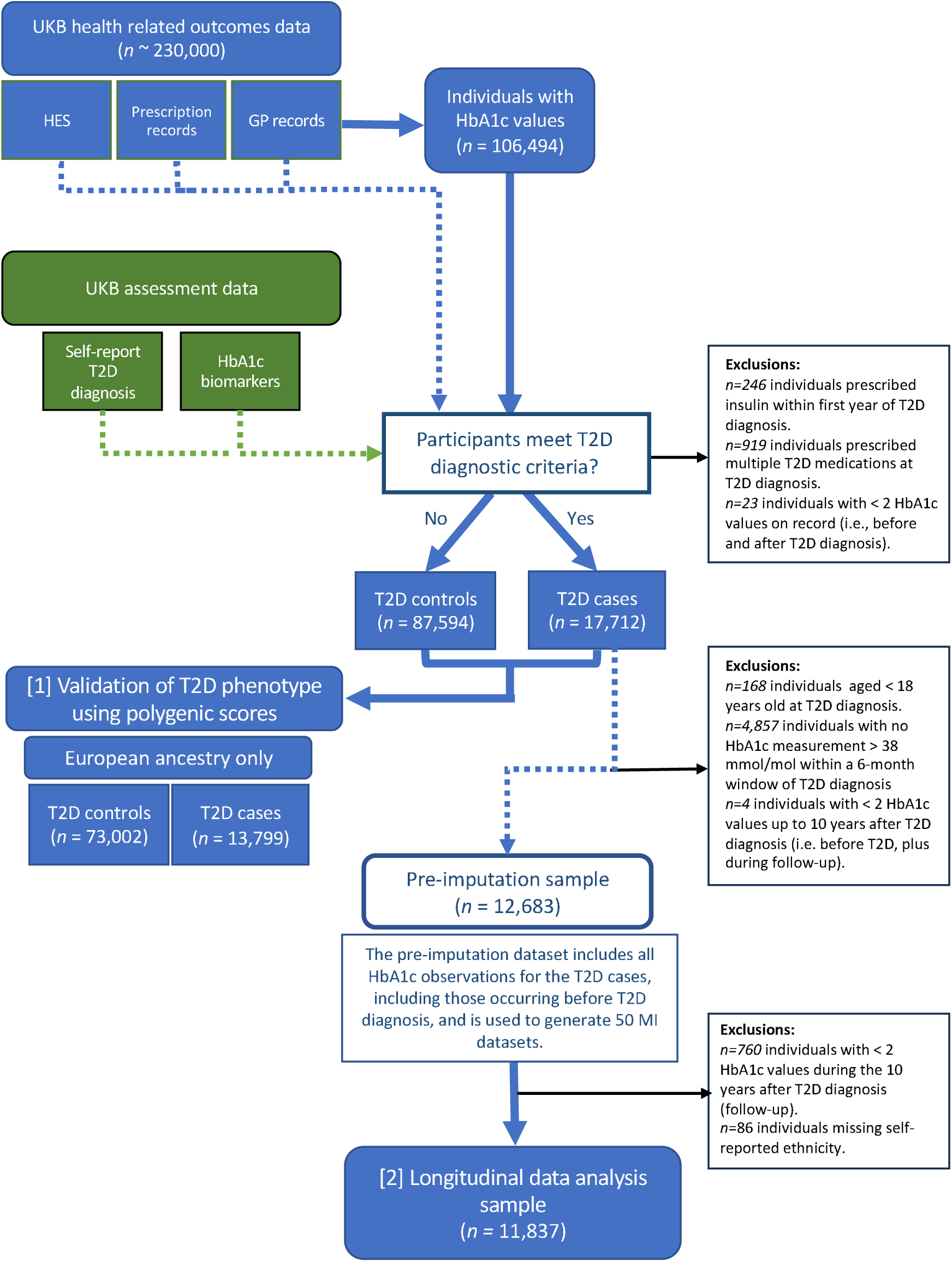
Flow diagram of UK Biobank (UKB) participant selection. T2D = type 2 diabetes. MDD = major depressive disorder. HES = hospital episode statistics. HbA1c = glycated haemaglobin. Validation of T2D diagnosis sample = T2D case and controls of European ancestry, meeting eligibility criteria outlined in Methods, including individual-level genetic analysis inclusion criteria (described in SuppMethods 1). Imputed datasets used for analyses (2) and (3) (adjusted mixed effects model for HbA1c over time and within-individual HbA1c variation).

**Figure 2:**
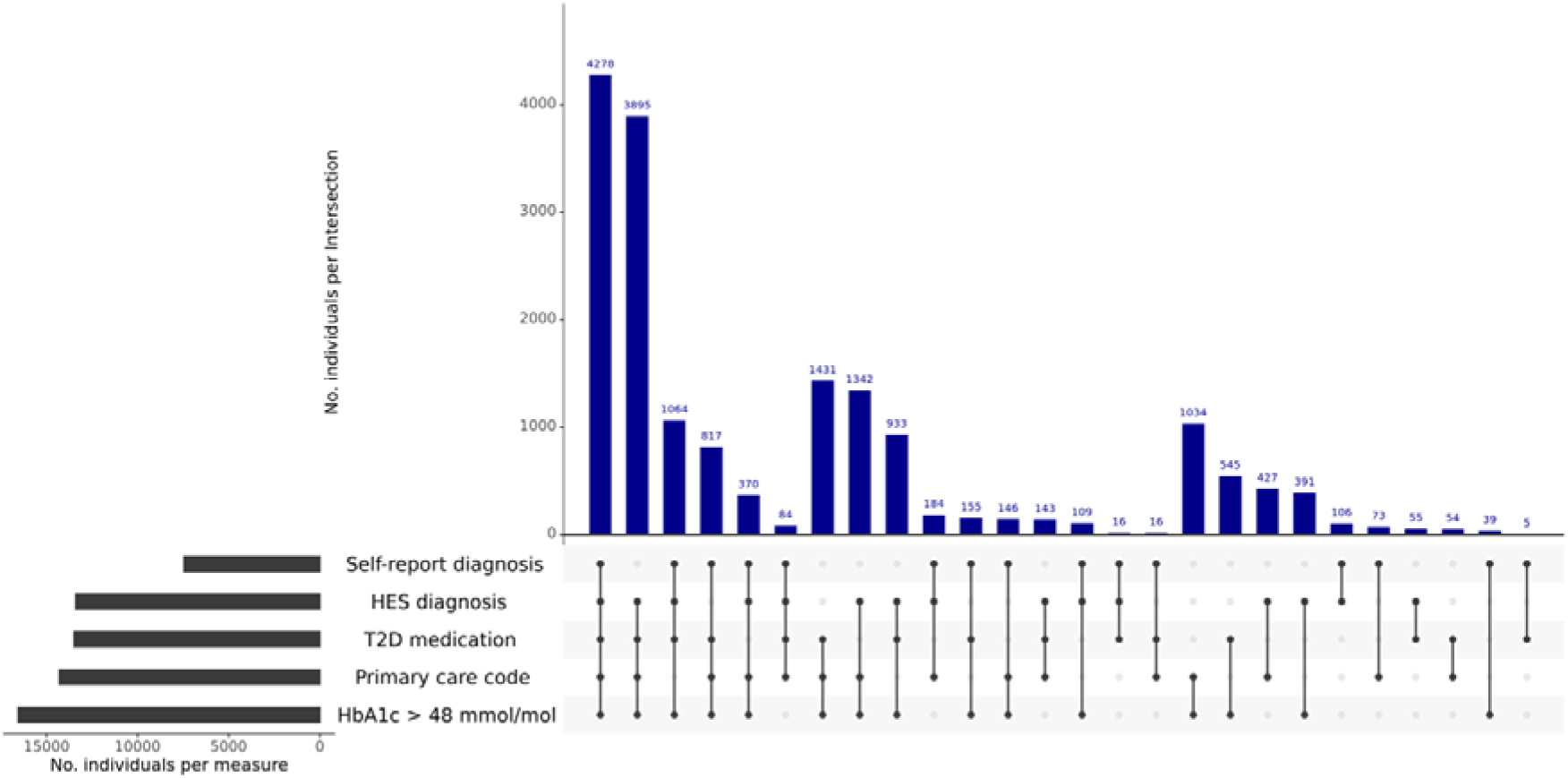
Contributions of each input to T2D phenotype definition Horizontal bars indicate the number of individuals who met criteria for the corresponding T2D phenotypes. Vertical bars indicate the number of individuals endorsing combinations of the five T2D phenotypes.

### Outcome measures

For the T2D diagnostic criteria validation, the outcome was T2D case-control status for UKB participants of European ancestry with primary care records available. For the longitudinal analysis, the outcome was repeated measures of HbA1c (mmol/mol) after T2D diagnosis.

HbA1c data were taken from: 1) primary care records up to 2017 (SuppTable S4), where older observations recorded in %-units were converted to mmol/mol^17^, and 2) all UKB biomarker assessments (2006-2016), where a validated correction was applied to account for lower average HbA1c values from the UKB biomarker panel compared to primary care^18^ (SuppMethods 4). The indexing date was T2D diagnosis, with a maximum follow-up period of 10 years.

### Exposures

In the longitudinal analysis, four MDD exposure variables were considered simultaneously. Two were related to individuals diagnosed with MDD prior to T2D (pre-T2D MDD), and two were related to individuals diagnosed with MDD after their T2D diagnosis (post-T2D MDD). *Pre-T2D MDD exposures:* (1) History of MDD at T2D diagnosis (MDD_index). This binary variable indicates whether an individual had ever received a MDD diagnosis at the index date. (2) Pre-T2D MDD duration at index (years). This semi-continuous variable quantifies the time between MDD diagnosis and T2D diagnosis when MDD_index equals 1, and is 0 otherwise. These time-invariant exposures were used to examine the impact of pre-T2D MDD on HbA1c, including interactions with T2D disease duration.

*Post-T2D MDD exposures:* (1) Change in MDD diagnosis (MDD_change). A time-varying binary variable indicating, at each observation time (*t*), whether an individual has been diagnosed with MDD between *t* and the index date. It captures information about individuals diagnosed with MDD during follow-up. For individuals with no MDD diagnosis occurring before *t*, and those diagnosed prior to T2D diagnosis, this variable is set to 0. (2) Post-T2D MDD duration (years). A semi-continuous, time-varying covariate equalling the time between MDD diagnosis and *t* if MDD_change equals 1, and 0 otherwise. This variable allows post-T2D MDD participants to have different HbA1c time-slopes after their MDD diagnosis. Note, UKB participants classified as having a MDD diagnosis required at least two diagnostic codes for a depressive disorder or episode diagnosis in the linked primary care records^16^.

### Covariates

Covariates were extracted from UKB assessments and/or primary care data. Covariates extracted from UKB initial assessments were: sex, assessment centre, self-reported ethnicity, Townend Deprivation Index (TDI), qualifications, ever smoked, and never consumed alcohol. Covariates extracted from both UKB assessments and primary care data were: age at T2D diagnosis, HbA1c at T2D diagnosis, body mass index (BMI) at T2D diagnosis (SuppMethods 6), systolic (SBP) and diastolic (DBP) blood pressure at T2D diagnosis (SuppMethods 6), number of HbA1c, BMI and blood pressure measurements taken prior to T2D diagnosis, and T2D disease duration (time). Covariates extracted from primary care only were diabetic medications. Glucose lowering medication at each HbA1c observation was identified using prescription records up to three months prior to HbA1c measurement (SuppMethods 2). This information was grouped into four medication categories: 1) M0 (‘no medication’), 2) M1 (‘metformin or a single medication’/ monotherapy), 3) M2 (‘two medications’/ dual-therapy), and 4) M3 (either ‘3 or more medications’ or ‘insulin’). Two medication variables were then created. Firstly, a binary variable indicating monotherapy at T2D diagnosis versus no prescribed T2D medication (M0 vs M1 at diagnosis). Secondly, a time-varying medication variable using the four categories M0–M3.

With the exception of time and time-varying medication, covariates were treated as baseline measurements. However, it is important to note that the index date and UKB assessment dates are different. For example, measurements from UKB initial assessments (TDI, qualifications, etc) were collected between 2006 and 2010, and 56% of individuals were diagnosed outside of this time-frame. However, results from models with and without UKB initial assessment covariates yielded similar conclusions (SuppTable S10). Further details on covariates are available in SuppTable 5A.

### Statistical analysis

All analyses were performed using R version 4.2.2, and visualised using ggplot2. *Validation of T2D diagnostic criteria:* To validate the T2D definition, we tested whether PGSs for T2D^19^ predicted T2D case-control status. PGSs, calculated using PRSice v2^20,21^ at eleven *P*-value thresholds, were tested for association with T2D case-control status, adjusting for six genetic ancestry principal components, assessment centre and genetic batch effect (SuppMethods 7).

*Longitudinal analysis:* We employed linear mixed effects models (MEMs) to investigate longitudinal associations between HbA1c and MDD using the nlme package^22,23^. All models incorporated random intercepts and time slopes, with a continuous-time autoregressive 1 (CAR1) residual correlation structure to account for autocorrelation. A series of three analyses were performed.

**(1) Unadjusted model with selection**. This analysis focused on the primary fixed effects of time and the MDD exposures. Time (T2D disease duration) was modelled using a restricted cubic spline (RCS) with four knots to allow for a non-linear temporal trend in mean HbA1c ^24,25^. Interactions between the pre-T2D MDD exposures and time-splines were considered, with the impact of post-T2D MDD on HbA1c time-slopes captured by post-T2D MDD duration. Selection of the final model was determined by Akaike Information Criterion (AIC) and likelihood ratio tests (LRTs). Models with semi-continuous exposures required their corresponding binary indicator to be included.
**(2) Adjusted model.** The selected unadjusted model was extended to include covariates, plus interactions between the time-splines and HbA1c at T2D diagnosis, BMI at T2D diagnosis and the medication variables.
**(3) Residual within-subject variation in HbA1c.** To explore the association between within-subject variation and MDD, the adjusted model was updated to allow residual variation to differ by MDD diagnosis variables. Firstly, we assessed if pre-T2D MDD (MDD_index) individuals had different residual variation compared to other participants. Secondly, we used MDD_change to investigate if post-T2D MDD individuals had different residual variation after their MDD diagnosis. Thirdly, a restrospective, time-invariant binary indicator for post-T2D MDD individuals was introduced to assess any differences in within-subject variation over all follow-up. Likelihood ratio tests (LRTs) were used to test for MDD diagnosis-related heterogeneity in residual variation.

HbA1c at T2D diagnosis had a high level of missingness (40%), but all included participants had a HbA1c measurement within a six-month window of this date. Multiple imputation (MI) with the Amelia package^26^ generated 50 MI datasets (SuppMethods 8) that were used in analyses (2) and (3). MEM estimates were pooled using Rubins method, with pooled Wald-like *P*-values presented for the fixed effects^27,28^. Parameter estimates for RCSs are difficult to interpret. Therefore, in addition to a summary of the MDD exposure fixed effects (including p-values from t-tests), results are presented using plots of predicted HbA1c^29,30^. Details of covariates included in MI and/ or MEMs are given in SuppTables S5A and 5B.

## Results

### Validation of T2D diagnostic criteria

The eligible UKB sample consisted of 17,712 individuals with T2D (Figure 2), with 24% of individuals having evidence of T2D from all five data sources considered. Participants with T2D tend to be male (61%) with an average age at T2D diagnosis of 57 years (IQR: 51-64 years). Applying genetic quality control criteria to participants of European ancestry provided 13,799 T2D cases and 73,002 controls. T2D PGSs were significantly associated with T2D case-control status at all PGS *P*-value thresholds and explained up to 2.8% of T2D liability (SuppTable S6).

### Longitudinal analysis

In total, 11,837 T2D cases met inclusion criteria for longitudinal analysis. Median follow-up time was 6.9 years (IQR: 3.5–9.3 years). Table 1 presents sample characteristics stratified by MDD subgroup: ‘no MDD’ (89% without an MDD diagnosis), ‘pre-T2D MDD’ (9% with MDD diagnosed prior to T2D) and ‘post-T2D MDD’ (2% diagnosed with MDD during the 10 year follow-up). Groups with a diagnosis of MDD include a higher proportion of females and have higher median TDI, which is particularly notable in the post-T2D MDD group. Additionally, the post-T2D MDD group had an earlier median age at T2D diagnosis, leading to a longer median follow-up time. This group also had the highest proportion of patients in medication group M3 (insulin and/or three or more T2D medications) by the end of follow-up. A higher proportion of the post-T2D MDD group are missing HbA1c at diagnosis (52% compared to 43% for the pre-T2D MDD group and 42% for the no MDD group).

**Table 1.**
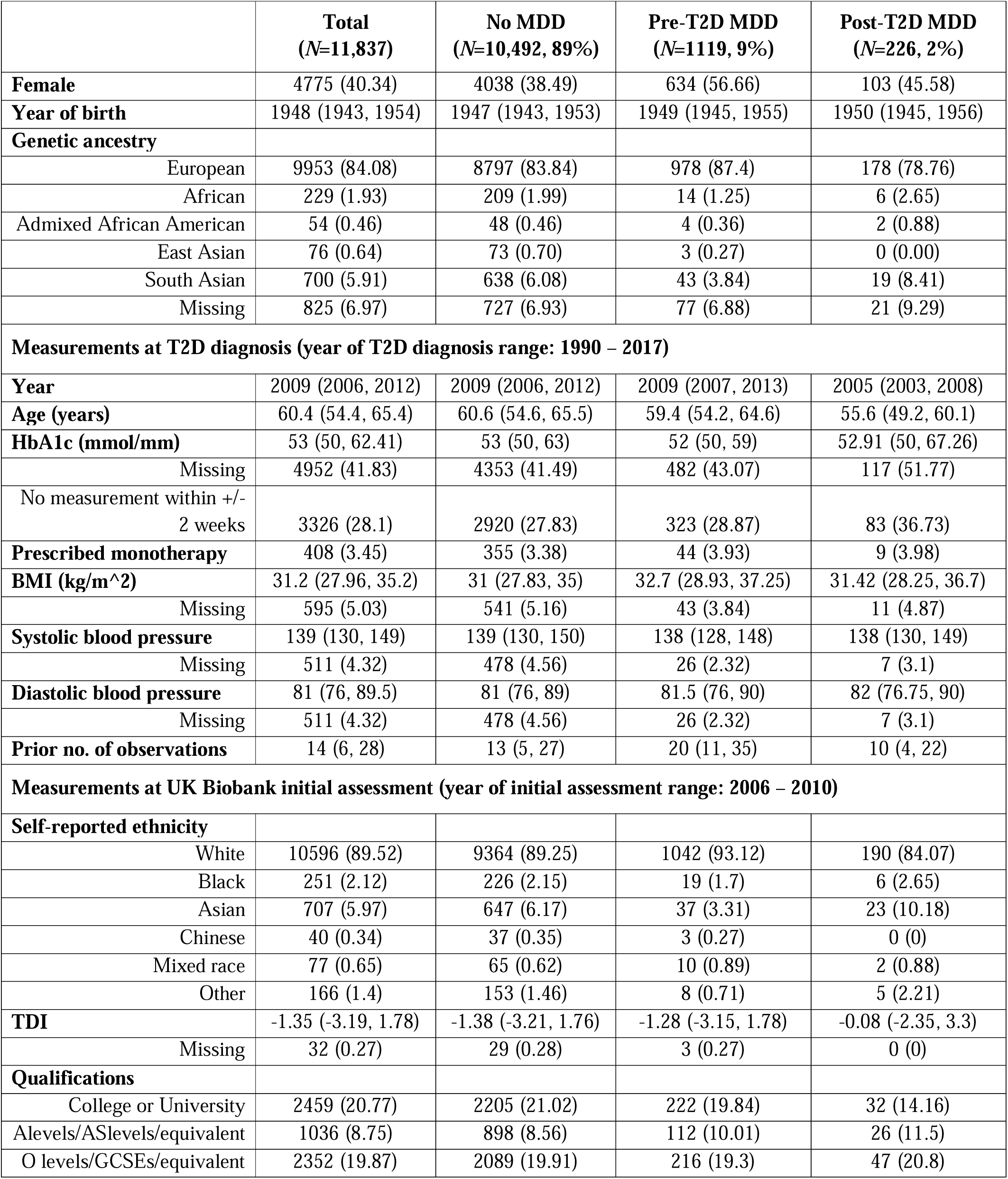
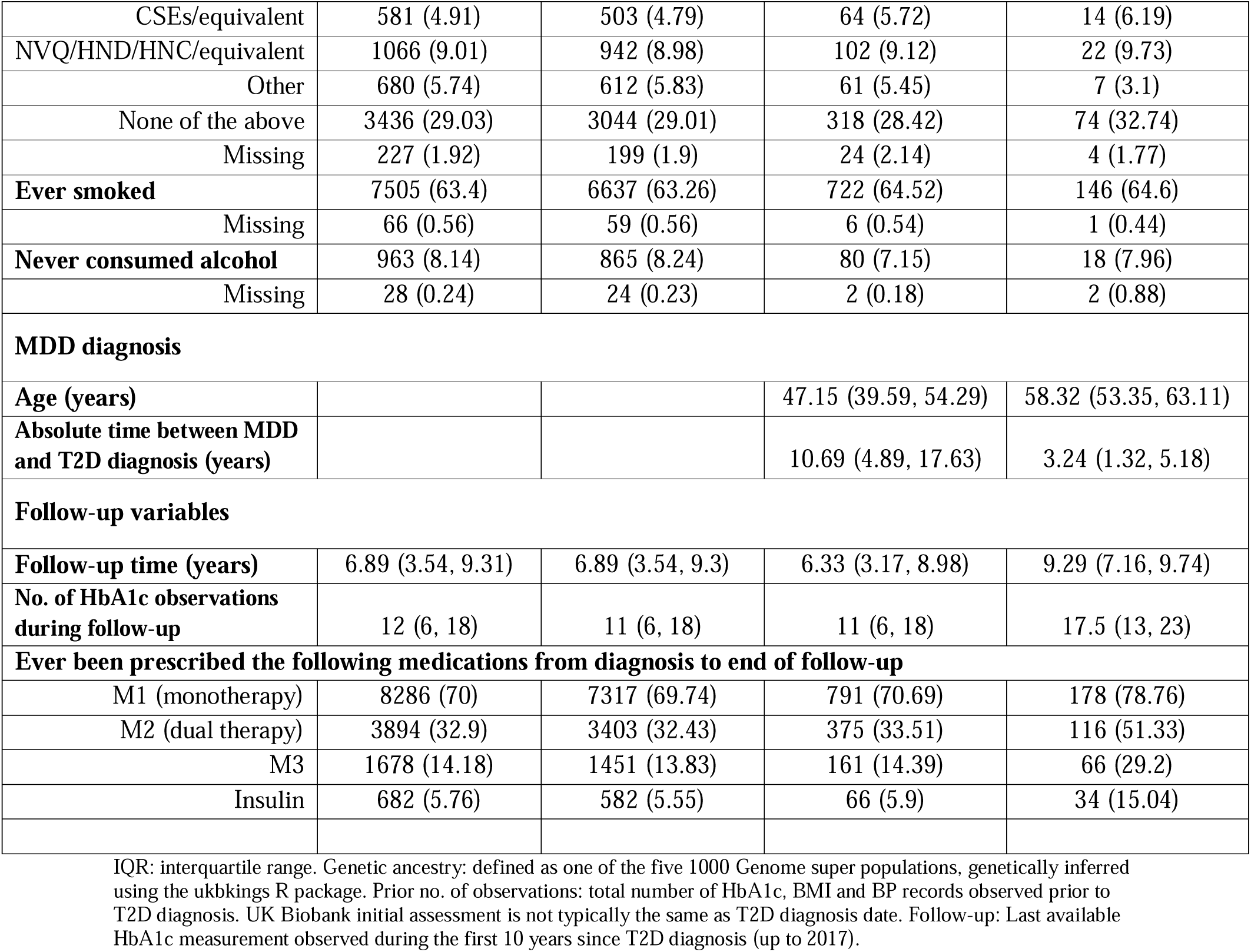
Characteristics of the longitudinal analysis study participants at T2D diagnosis, stratified by MDD subgroups. Columns present N (%) for categorical variables and median (IQR) for continuous variables.

|  | **Total**<br>**(N=11,837)** | **No MDD**<br>**(N=10,492, 89%)** | **Pre-T2D MDD**<br>**(N=1119, 9%)** | **Post-T2D MDD**<br>**(N=226, 2%)** |
| --- | --- | --- | --- | --- |
| **Female** | 4775 (40.34) | 4038 (38.49) | 634 (56.66) | 103 (45.58) |
| **Year of birth** | 1948 (1943, 1954) | 1947 (1943, 1953) | 1949 (1945, 1955) | 1950 (1945, 1956) |
| **Genetic ancestry** |  |  |  |  |
| European | 9953 (84.08) | 8797 (83.84) | 978 (87.4) | 178 (78.76) |
| African | 229 (1.93) | 209 (1.99) | 14 (1.25) | 6 (2.65) |
| Admixed African American | 54 (0.46) | 48 (0.46) | 4 (0.36) | 2 (0.88) |
| East Asian | 76 (0.64) | 73 (0.70) | 3 (0.27) | 0 (0.00) |
| South Asian | 700 (5.91) | 638 (6.08) | 43 (3.84) | 19 (8.41) |
| Missing | 825 (6.97) | 727 (6.93) | 77 (6.88) | 21 (9.29) |
| **Measurements at T2D diagnosis (year of T2D diagnosis range: 1990 – 2017)** |  |  |  |  |
| **Year** | 2009 (2006, 2012) | 2009 (2006, 2012) | 2009 (2007, 2013) | 2005 (2003, 2008) |
| **Age (years)** | 60.4 (54.4, 65.4) | 60.6 (54.6, 65.5) | 59.4 (54.2, 64.6) | 55.6 (49.2, 60.1) |
| **HbA1c (mmol/mm)** | 53 (50, 62.41) | 53 (50, 63) | 52 (50, 59) | 52.91 (50, 67.26) |
| Missing | 4952 (41.83) | 4353 (41.49) | 482 (43.07) | 117 (51.77) |
| No measurement within +/- 2 weeks | 3326 (28.1) | 2920 (27.83) | 323 (28.87) | 83 (36.73) |
| **Prescribed monotherapy** | 408 (3.45) | 355 (3.38) | 44 (3.93) | 9 (3.98) |
| **BMI (kg/m<sup>2</sup>)** | 31.2 (27.96, 35.2) | 31 (27.83, 35) | 32.7 (28.93, 37.25) | 31.42 (28.25, 36.7) |
| Missing | 595 (5.03) | 541 (5.16) | 43 (3.84) | 11 (4.87) |
| **Systolic blood pressure** | 139 (130, 149) | 139 (130, 150) | 138 (128, 148) | 138 (130, 149) |
| Missing | 511 (4.32) | 478 (4.56) | 26 (2.32) | 7 (3.1) |
| **Diastolic blood pressure** | 81 (76, 89.5) | 81 (76, 89) | 81.5 (76, 90) | 82 (76.75, 90) |
| Missing | 511 (4.32) | 478 (4.56) | 26 (2.32) | 7 (3.1) |
| **Prior no. of observations** | 14 (6, 28) | 13 (5, 27) | 20 (11, 35) | 10 (4, 22) |
| **Measurements at UK Biobank initial assessment (year of initial assessment range: 2006 – 2010)** |  |  |  |  |
| **Self-reported ethnicity** |  |  |  |  |
| White | 10596 (89.52) | 9364 (89.25) | 1042 (93.12) | 190 (84.07) |
| Black | 251 (2.12) | 226 (2.15) | 19 (1.7) | 6 (2.65) |
| Asian | 707 (5.97) | 647 (6.17) | 37 (3.31) | 23 (10.18) |
| Chinese | 40 (0.34) | 37 (0.35) | 3 (0.27) | 0 (0) |
| Mixed race | 77 (0.65) | 65 (0.62) | 10 (0.89) | 2 (0.88) |
| Other | 166 (1.4) | 153 (1.46) | 8 (0.71) | 5 (2.21) |
| **TDI** | -1.35 (-3.19, 1.78) | -1.38 (-3.21, 1.76) | -1.28 (-3.15, 1.78) | -0.08 (-2.35, 3.3) |
| Missing | 32 (0.27) | 29 (0.28) | 3 (0.27) | 0 (0) |
| **Qualifications** |  |  |  |  |
| College or University | 2459 (20.77) | 2205 (21.02) | 222 (19.84) | 32 (14.16) |
| A levels/AS levels/equivalent | 1036 (8.75) | 898 (8.56) | 112 (10.01) | 26 (11.5) |
| O levels/GCSEs/equivalent | 2352 (19.87) | 2089 (19.91) | 216 (19.3) | 47 (20.8) |
| CSEs/equivalent | 581 (4.91) | 503 (4.79) | 64 (5.72) | 14 (6.19) |
| NVQ/HND/HNC/equivalent | 1066 (9.01) | 942 (8.98) | 102 (9.12) | 22 (9.73) |
| Other | 680 (5.74) | 612 (5.83) | 61 (5.45) | 7 (3.1) |
| None of the above | 3436 (29.03) | 3044 (29.01) | 318 (28.42) | 74 (32.74) |
| Missing | 227 (1.92) | 199 (1.9) | 24 (2.14) | 4 (1.77) |
| **Ever smoked** | 7505 (63.4) | 6637 (63.26) | 722 (64.52) | 146 (64.6) |
| Missing | 66 (0.56) | 59 (0.56) | 6 (0.54) | 1 (0.44) |
| **Never consumed alcohol** | 963 (8.14) | 865 (8.24) | 80 (7.15) | 18 (7.96) |
| Missing | 28 (0.24) | 24 (0.23) | 2 (0.18) | 2 (0.88) |
| **MDD diagnosis** |  |  |  |  |
| **Age (years)** |  |  | 47.15 (39.59, 54.29) | 58.32 (53.35, 63.11) |
| **Absolute time between MDD and T2D diagnosis (years)** |  |  | 10.69 (4.89, 17.63) | 3.24 (1.32, 5.18) |
| **Follow-up variables** |  |  |  |  |
| **Follow-up time (years)** | 6.89 (3.54, 9.31) | 6.89 (3.54, 9.3) | 6.33 (3.17, 8.98) | 9.29 (7.16, 9.74) |
| **No. of HbA1c observations during follow-up** | 12 (6, 18) | 11 (6, 18) | 11 (6, 18) | 17.5 (13, 23) |
| **Ever been prescribed the following medications from diagnosis to end of follow-up** |  |  |  |  |
| M1 (monotherapy) | 8286 (70) | 7317 (69.74) | 791 (70.69) | 178 (78.76) |
| M2 (dual therapy) | 3894 (32.9) | 3403 (32.43) | 375 (33.51) | 116 (51.33) |
| M3 | 1678 (14.18) | 1451 (13.83) | 161 (14.39) | 66 (29.2) |
| Insulin | 682 (5.76) | 582 (5.55) | 66 (5.9) | 34 (15.04) |

IQR: interquartile range. Genetic ancestry: defined as one of the five 1000 Genome super populations, genetically inferred using the ukbkings R package. Prior no. of observations: total number of HbA1c, BMI and BP records observed prior to T2D diagnosis. UK Biobank initial assessment is not typically the same as T2D diagnosis date. Follow-up: Last available HbA1c measurement observed during the first 10 years since T2D diagnosis (up to 2017).

#### Unadjusted model

The selected unadjusted model included time-splines and the four MDD exposures, with no significant interaction between the pre-T2D MDD exposures and time (*p* = 0.3072; Table 2). This implies the difference in mean HbA1c between pre-T2D MDD individuals and individuals without MDD depends on the time between MDD and T2D diagnoses, the effect of which is constant over T2D disease duration. Having a history of MDD prior to T2D is associated with a 0.92 mmol/mol increase (95% CI: [0.00, 1.84]) in HbA1c, but earlier onset of pre-T2D MDD is associated with lower HbA1c (Table 3). To illustrate this, Figure 3 plots predicted HbA1c (mmol/mol) over time for four example individuals with either no MDD or with MDD diagnosed 2.2 years, 10.7 years or 27.5 years before T2D (10^th^, 50^th^ and 90^th^ percentile of pre-T2D MDD duration at index respectively). Differences in HbA1c between these three pre-T2D MDD individuals and the no MDD individual at any point during follow-up are 0.72 (95% CI: [-0.11, 1.55]), -0.06 (95% CI: [-0.65, 0.54]) and -1.60 (95 CI: [-2.63, -0.57]) for the 10^th^, 50^th^ and 90^th^ percentile of pre-T2D MDD duration respectively. Those diagnosed with MDD ∼10 years before T2D have HbA1c levels over time equivalent to those without MDD, with those diagnosed earlier having lower HbA1c and those diagnosed closer to their T2D diagnosis having higher HbA1c. Our example shows that only individuals diagnosed with MDD many decades before T2D have a 95% CI for difference in HbA1c relative to no MDD that excludes 0 (SuppTable S8a).

**Figure 3.**
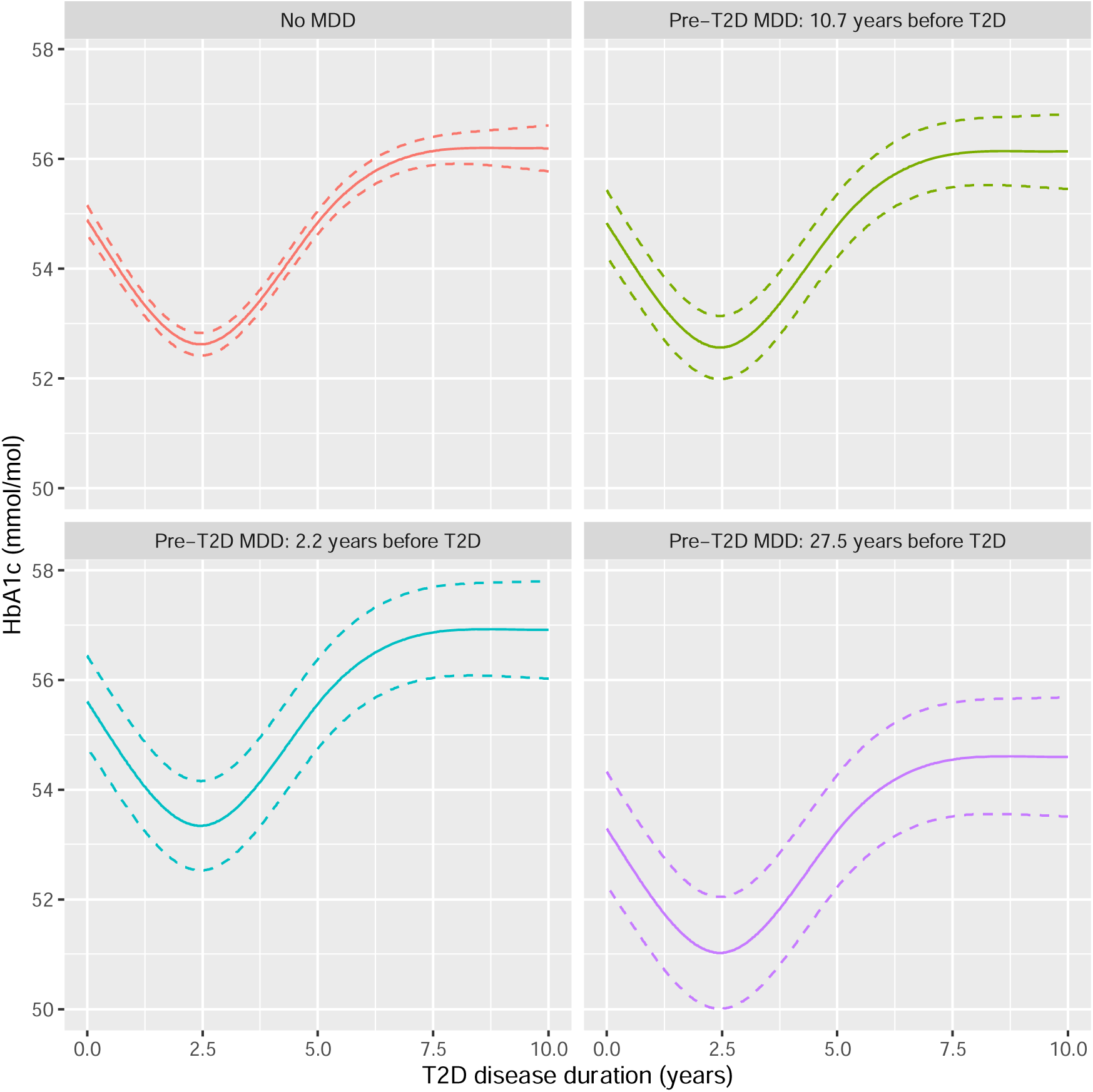
Predicted HbA1c (mmol/mol) over T2D disease duration (years) from the unadjusted model, for four example individuals: no MDD and three pre-T2D MDD individuals diagnosed with MDD 2.2 (10 percentile), 10.7 (median) and 27.5 (90^th^ percentile) years before T2D.

**Table 2.**
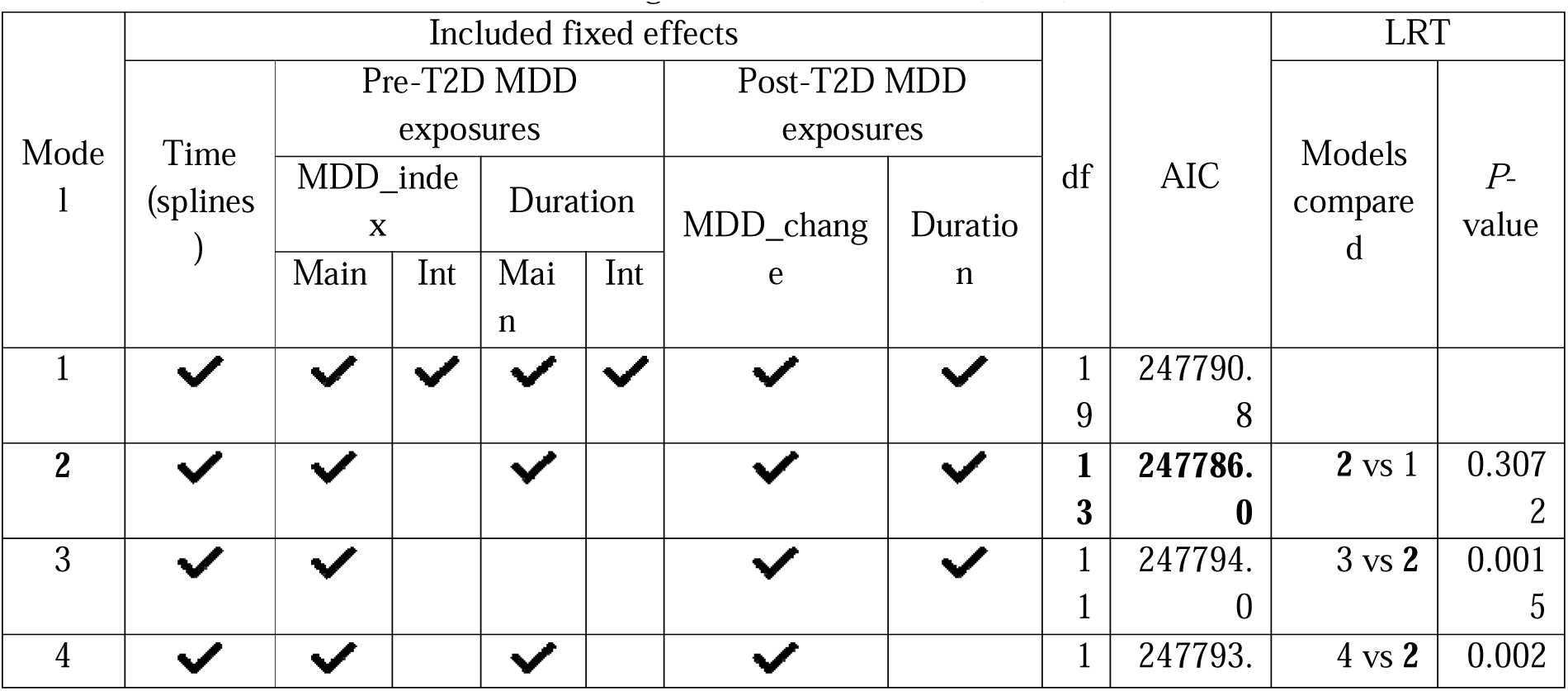
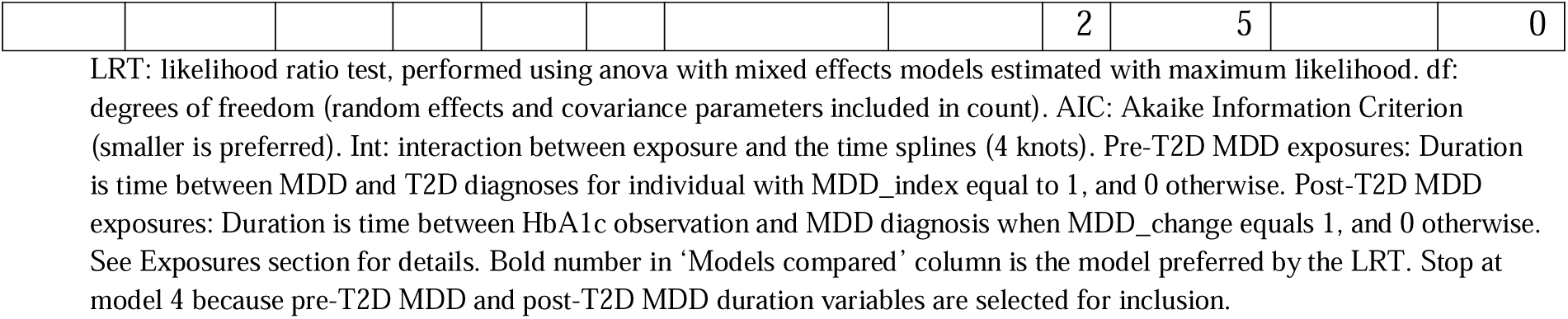
Unadjusted model selection summary. Model 2, with mean HbA1c being a function of time (4 knots splines for T2D disease duration), the pre-T2D MDD exposures and the post-T2D exposures, has the lowest AIC and is selected using likelihood ratio tests (LRTs).

**Table 3.** Fixed effect estimates for MDD exposure variables from the unadjusted and pooled adjusted linear mixed effects models.

| MDD Exposure Variables | Unadjusted model |  |  | Pooled adjusted model |  |  |
| --- | --- | --- | --- | --- | --- | --- |
|  | Beta | 95% CI | P-value | Beta | 95% CI | P-value |
| **Pre-T2D MDD exposure variables** |  |  |  |  |  |  |
| Pre-T2D MDD | 0.92 | ( 0.00, 1.84) | 0.0496 | 0.84 | (-0.06, 1.73) | 0.0678 |
| Pre-T2D MDD duration to index (years) | -0.09 | (-0.15, -0.04) | 0.0015 | -0.04 | (-0.09, 0.02) | 0.1637 |
| **Post-T2D MDD exposure variables** |  |  |  |  |  |  |
| Post-T2D MDD | -0.50 | (-1.63, 0.64) | 0.3911 | -0.63 | (-1.73, 0.46) | 0.2555 |
| Post-T2D MDD duration (years) | 0.53 | ( 0.19, 0.87) | 0.0020 | 0.51 | ( 0.17, 0.86) | 0.0038 |

The unadjusted model contains the MDD exposure variables and T2D disease durations splines as fixed effects. The adjusted model includes the following additional fixed effects: sex, UKB assessment centre, age at T2D diagnosis, BMI at T2D diagnosis, SBP and DBP at T2D diagnosis, HbA1c at T2D diagnosis, self-reported ethnicity, qualifications at UKB initial assessment, number of observations prior to T2D (BMI, BP and HbA1c), ever smoke, never consumed alcohol, T2D medications and interactions between T2D diagnosis and HbA1c at T2D, T2D medications and BMI. Presented p-values are from t-tests. Model parameters and p-values from the adjusted model were pooled from 50 MI datasets. SuppTabs S7 and S9 present full model results for the unadjusted and pooled adjusted model respectively.

For post-T2D MDD individuals, the unadjusted model included a small, non-significant decrease in HbA1c of -0.50 mmol/mol (95% CI: [-1.63, 0.64]) upon MDD diagnosis (Table 3). This initial MDD diagnosis effect is retained in the model due to the significant semi-continuous post-T2D MDD duration variable, capturing time since post-T2D MDD diagnosis (Table 3). For two post-T2D MDD individuals where MDD is diagnosed one year apart, HbA1c is expected to be 0.53 mmol/mol higher (95% CI: [0.19, 0.87]) in the individual diagnosed with MDD one year earlier (Table 3).

Figure 4 presents predicted HbA1c over time for an individual without MDD and three example individuals diagnosed with MDD 1 year, 3.8 years and 7.1 years after T2D (10^th^, 50^th^ and 90^th^ percentile respectively). The smaller sample size for the post-T2D MDD group is reflected in wider 95% CI after MDD diagnosis. An initial decrease in HbA1c at MDD diagnosis is followed by a steeper increase in HbA1c, so that a post-MDD individual will have a higher HbA1c three years after MDD diagnoisis compared to an individual with no MDD diagnosis (i.e. the 95% CI for the difference between predicted HbA1c for a post-T2D MDD individual and an individual without MDD is expected to exclude 0 after three years). Indivduals diagnosed with MDD earlier in their follow-up are expected to have higher HbA1c levels after a 10-year T2D disease duration compared to those with later onset MDD or those without MDD diagnoses (SuppTable S8b). For example, at the end of the 10-year follow-up, an individual diagnosed with MDD one year after T2D is predicted to have a HbA1c of 60.49 (95% CI: [57.83, 63.15]) compared to 56.19 (95% CI: [55.78, 56.61]) for those without MDD.

**Figure 4.**
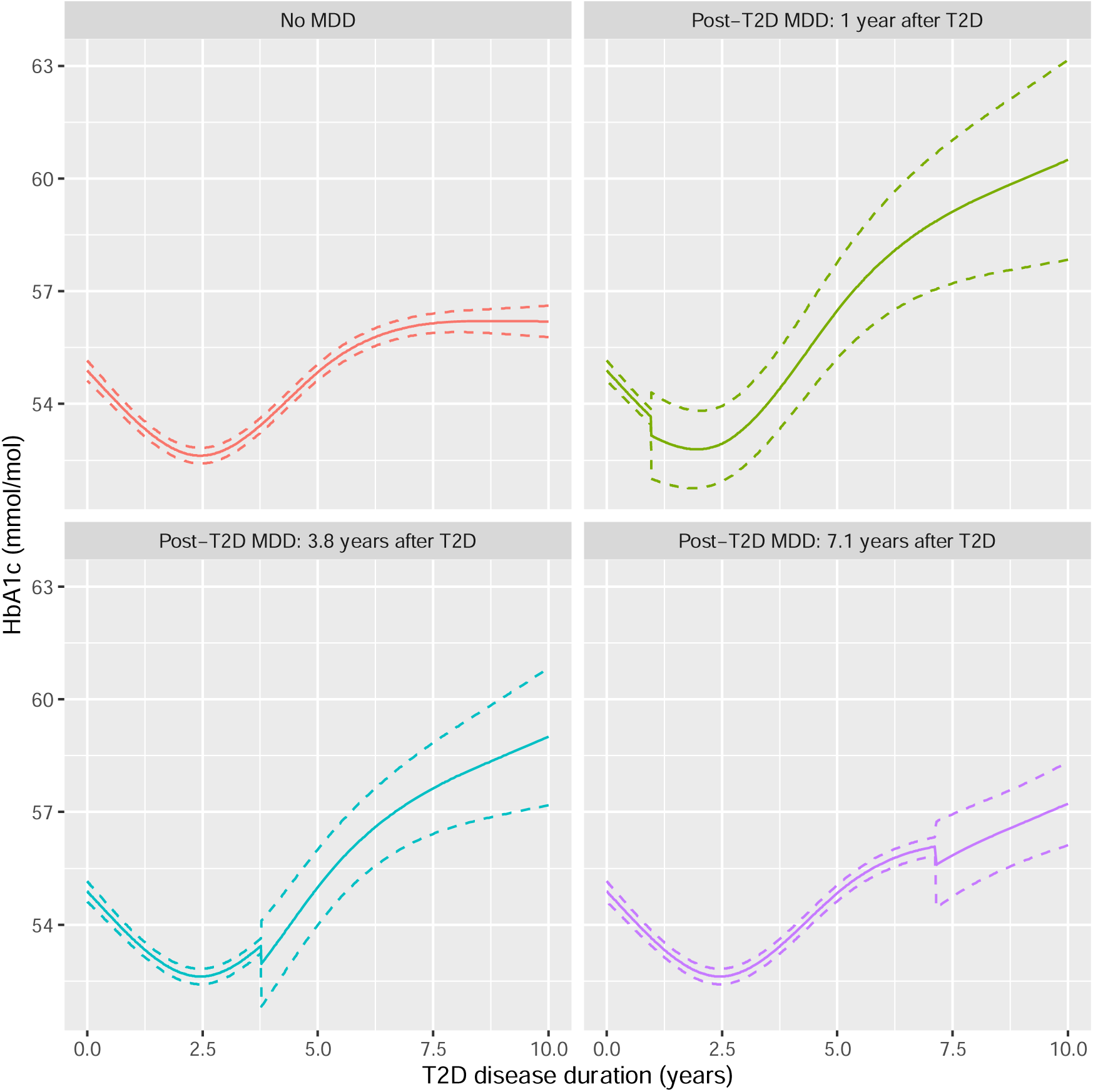
Predicted HbA1c (mmol/mol) over T2D disease duration (years) from the unadjusted model, for four example individuals: no MDD and three post-T2D MDD individuals diagnosed with MDD 1 (10 percentile), 3.8 (median) and 7.1 (90 percentile) years after T2D.

#### Adjusted model

After adjustment for all covariates, the absolute effect size for all MDD exposure variables are reduced (Table 3). In particular, after adjustment, the pre-T2D MDD duration effect size is no longer significant (*p* = 0.1637) suggesting that, given the included covariates, a history of MDD prior to T2D has no effect on HbA1c trends over T2D disease duration.

The pooled results from the adjusted model are similar to those from the unadjusted model for the post-T2D MDD exposures. The adjusted model suggests a small decrease in HbA1c after a diagnosis with MDD during follow-up (post-T2D MDD). As in the unadjusted results, this effect is non-significant (*p* = 0.2555). The adjusted effect size for post-T2D MDD duration is 0.51 (95% CI: [0.17, 0.86]), showing that earlier MDD diagnosis during follow-up is still associated with higher HbA1c. Full model summaries can be found in SuppTable S7 and S9 for the unadjusted and adjusted model respectively.

#### Within-patient HbA1c variability

Non-convergence was common for models allowing residual variation to differ by MDD_index (24%) and MDD_change (48%). Pooled results from converged models showed that within-patient variability in HbA1c was approximately the same when comparing pre-T2D MDD individuals (using MDD_index) to all others, and post-T2D MDD (using MDD_change) to all others (Table 4). However, when looking retrospectively for post-T2D MDD individuals (i.e. looking across all follow-up rather than only after MDD diagnosis), we found that within-patient HbA1c variation was 1.16 times higher (95% CI: [1.13. 1.19]) for the post-T2D MDD group compared to no MDD and pre-T2D MDD. This suggests that differences in HbA1c for those who develop MDD after T2D, compared to those with no or pre-existing MDD, start prior to MDD diagnosis.

**Table 4:**
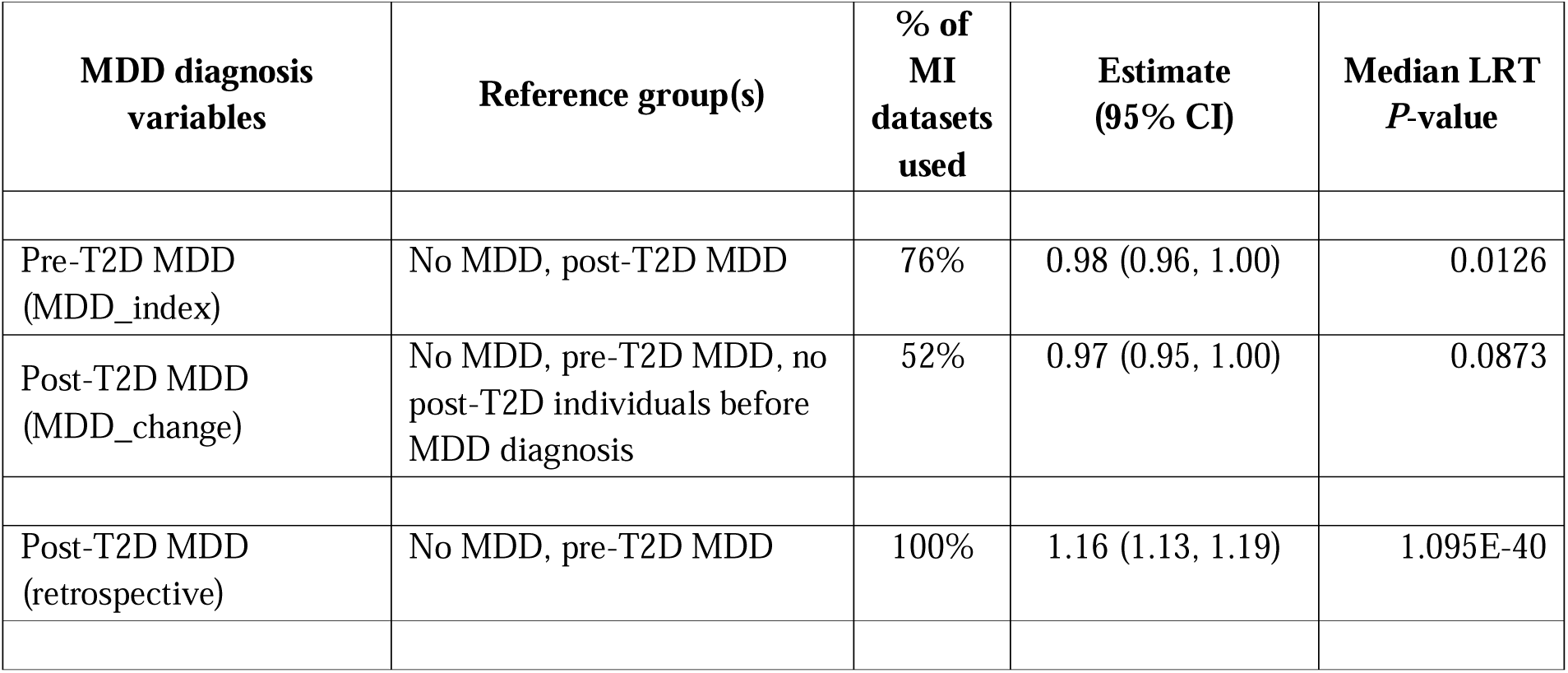
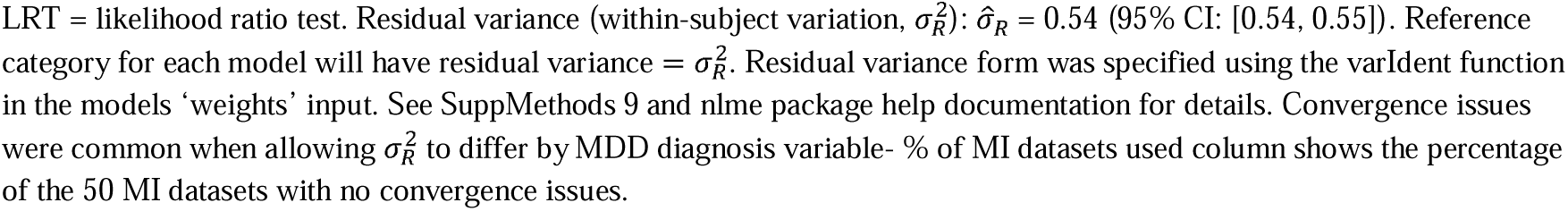
Pooled hypothesis testing for within-subject variability in HbA1c differing by MDD diagnosis variable. The parameter estimate presented represents the ratio of residual variation for MDD diagnosis versus reference level.

| MDD diagnosis variables | Reference group(s) | % of MI datasets used | Estimate (95% CI) | Median LRT P-value |
| --- | --- | --- | --- | --- |
| Pre-T2D MDD (MDD_index) | No MDD, post-T2D MDD | 76% | 0.98 (0.96, 1.00) | 0.0126 |
| Post-T2D MDD (MDD_change) | No MDD, pre-T2D MDD, no post-T2D individuals before MDD diagnosis | 52% | 0.97 (0.95, 1.00) | 0.0873 |
| Post-T2D MDD (retrospective) | No MDD, pre-T2D MDD | 100% | 1.16 (1.13, 1.19) | 1.095E-40 |

LRT = likelihood ratio test. Residual variance (within-subject variation, $\sigma_R^2$ ): $\hat{\sigma}_R = 0.54$ (95% CI: [0.54, 0.55]). Reference category for each model will have residual variance = $\sigma_R^2$ . Residual variance form was specified using the `varIdent` function in the models 'weights' input. See SuppMethods 9 and nlme package help documentation for details. Convergence issues were common when allowing $\sigma_R^2$ to differ by MDD diagnosis variable- % of MI datasets used column shows the percentage of the 50 MI datasets with no convergence issues.

## Discussion

This study tested the hypothesis that people with both T2D and MDD have poorer diabetic control over the course of T2D, as assessed by routine primary care monitoring of HbA1c, focusing on the role of timing and history of MDD diagnosis relative to T2D. We examined whether UKB participants with T2D and MDD have higher and more variable HbA1c levels over time compared to those with T2D alone. To do so, we utilised exposure variables that differentiated between individuals with a history of MDD before their T2D diagnosis (pre-T2D MDD) and those who received their MDD diagnosis after their T2D diagnosis (post-T2D MDD), while also incorporating time since MDD diagnosis.

For individuals with pre-T2D MDD, longitudinal modelling across 10 years of follow-up found that the time between MDD and T2D diagnoses was informative about HbA1c levels across T2D disease duration. Specifically, individuals diagnosed with MDD decades prior to T2D had lower HbA1c over time compared to individuals without MDD and those diagnosed closer to their T2D diagnosis date. After adjusting for covariates, the pre-T2D MDD variables became non-significant, which may be partly ascribed to the correlation between the time gap from pre-T2D MDD diagnosis to T2D and the age at T2D diagnosis (Spearman’s p = 0.23). Here, a longer gap between MDD and T2D diagnoses correlates with a later age at T2D diagnosis (SuppTable S11). However, the non-significance of pre-T2D MDD exposures after covariate adjustment may also be partly attributed to mediation rather than confounding. For example, increased numbers of pre-T2D measurements for blood pressure, BMI and HbA1c (a proxy for history of healthcare utilisation) were associated with lower HbA1c at T2D diagnosis (SuppTable S12). Given that an earlier diagnosis with MDD relative to T2D leads to more pre-T2D measurements (SuppTable S11), it is plausible that mediation is partially responsible for the loss of significance for pre-T2D MDD duration, through increased contact with healthcare professionals. Further work on the impact of pre-T2D MDD is therefore warranted, with extension to additional features of MDD beyond timing of initial diagnosis (e.g. most recent episode, anti-depressant prescriptions and number of episodes).

For individuals diagnosed with MDD after T2D, both the unadjusted and adjusted models found that time since MDD diagnosis was important in shaping the trajectory of HbA1c. The earlier the diagnosis of MDD during follow-up, the greater the expected difference in HbA1c between post-T2D MDD individuals and those without MDD at the end of follow-up. Post-T2D individuals also demonstrated some differences in within-patient HbA1c variation.

Results suggest that, given the model considered, after diagnosis with MDD during follow-up, within-patient variation does not differ from no MDD and pre-T2D MDD. However, if we look retrospectively across all follow-up and compare post-T2D MDD individuals to all others, we observe that within-patient HbA1c variation is 1.16 times higher for the post-T2D MDD individuals. This greater variability in HbA1c, driven by observations before MDD diagnosis, warrants further investigation to ensure appropriate public health and clinical advice is available. Post-T2D MDD individuals may indeed have higher HbA1c variability before MDD diagnosis. This is important because increased variability is associated with increased likelihood of adverse outcomes, including microvascular disease^31^. However, this result may also arise from mean HbA1c trends for post-T2D MDD individuals deviating from those with no or pre-existing MDD before their MDD diagnosis. Given that MDD diagnosis requires the presence of symptoms for two or more weeks, it is plausible that the impact of MDD symptoms on HbA1c begin before formal diagnosis. Additionally, diagnostic delays, as demonstrated in a study of primary care records in Spain (mean delay of 9.89 weeks)^32^ may be present. Therefore, exploring the feasibility of using HbA1c trends for MDD prediction in individuals with T2D could be an interesting avenue of research.

Previous work on mean HbA1c has provided inconclusive evidence for the impact of MDD^9^. Nevertheless, in line with our results, two larger studies did find associations between mean HbA1c over time and depression. A meta-analysis found a modest association between depressive symptoms and HbA1c (3-year mean follow-up)^9^, and a prospective study in veterans with T2D found that HbA1c was slightly higher for individuals with depression (5-year follow-up)^33^.

A previous study^10^ showed differences in T2D clinical characteristics by timing of MDD and T2D diagnoses, with a higher rate of diabetic complications in the post-T2D MDD group, but not the pre-TDD MDD group, compared to the no MDD group. Differences in glycaemic control between these MDD subgroups is a possible explanation for the observed clinical differences. The observed increase in HbA1c levels with time since MDD diagnosis for post-T2D MDD individuals, along with the retrospectively identified higher variability across follow-up, supports this hypothesis, demonstrating the potential importance of the relative timing of MDD onset.

Our analysis revealed a linear effect for time since post-T2D MDD diagnosis, meaning the impact of post-T2D MDD increases with years since MDD diagnosis. Due to the smaller sample size of the post-T2D MDD group (*n*=226), this study may have been underpowered to detect non-linear trends for this post-T2D MDD duration variable (SuppMethods 10). Future work using larger samples and different methods of capturing the time between T2D and MDD diagnoses (e.g. joint models for HbA1c over time and time-to-MDD diagnosis) may identify such trends, and determine if patients diagnosed with MDD after T2D require closer monitoring, or whether a specific time window is crucial for glycaemic control.

MDD episodes after T2D diagnosis are hypothesised to have a greater effect on glycaemic control, which we have confirmed by showing greater mean and within-subject variability in HbA1c levels for post-T2D MDD individuals. These individuals visited their GP after their T2D diagnosis and had a MDD code recorded. Multiple pathways, particularly behavioural, could explain this association. Several studies have shown that patients with both MDD and T2D have worse T2D self-management, are less able to keep the medical appointments, are less physically active and less able to adhere to dietary requirements, possibly leading to hyperglycaemia^34–36^. Our study highlights the importance of diagnosis timing, with possible need for targeted interventions based on clinical history.

Our study has several limitations. Firstly, primary care data are collected when patients visit their GP, and between-patient differences in this visiting process can bias results^37,38^. Individuals with T2D should see their GP every 3-6 months^11^ and differences may affect results. For example, less healthy individuals may interact with GPs more frequently and contribute more observations. The models used attempted to reduce this potential bias, as the CAR1 error structure ensured that observations measured closely in time provided less information than those taken far apart. However, T2D cases with MDD may miss more scheduled appointments, and our analysis does not account for this.

Secondly, as a retrospective observational study, confounding may bias results. For example, antidepressant medication can lead to weight gain^39^, which can negatively affect HbA1c levels^40^. While we adjusted for confounders such as BMI and blood pressure, these covariates were time invariant and the timing of measurement differed for each participant (SuppMethods 6). Future studies could consider time-varying confounding and include additional risk factors (e.g. antidepressant medications, cardiovascular conditions).

Thirdly, HbA1c at T2D diagnosis had a high level of missingness, which disproportionately affected individuals in the post-T2D MDD group, and highlights the challenges of working with real-world data. While we utilised MI, this approach has limitations and depends on the model selected and data availability. However, results for the post-T2D MDD exposures were similar in the unadjusted model (no MI required) and the adjusted model (MI used).

The validity of the age at MDD onset, determined using the date of the first MDD diagnostic code, presents a further limitation. For older individuals the validity of their MDD onset is unknown, given primary care records are only available after 1990, while mean age at diagnosis of MDD is around 30 years^41^. Replication in the Clinical Practice Research Datalink, which has GP records from 1987 and no age limits, would therefore be useful^42^.

## Conclusions

This study, utilising UKB primary care records, highlights the importance of considering the temporal relationships between T2D and MDD in the context of glycaemic control. Findings reveal a non-linear trend in HbA1c levels over time and demonstrated that within a 10-year window, those who develop MDD after T2D diagnosis had increased HbA1c levels and, prior to MDD diagnosis, greater variability, supporting the finding that individuals with both conditions have poorer health outcomes. Regular routine screening for depression, integrated mental health support and closer monitoring may reduce the adverse consequences associated with both T2D and MDD.

## Data Availability

Data is available from UK Biobank subject to standard access procedures (www.ukbiobank.ac.uk).

http://www.ukbiobank.com

## List of abbreviations

HbA1c: Glycated haemaglobin
T2D: Type 2 diabetes
MDD: Major depressive disorder
UKB: UK Biobank
GP: General practioner
PGS: Polygenic score
BMI: Body mass index
SBP: Systolic blood pressure
DBP: Diastolic blood pressure
MEM: Mixed effects model
CAR1: Continuous-time autoregressive 1
LRT: Likelihood ratio test
MI: Multiple imputation
CI: Confidence interval
IQR: Interquartile range
HES: Hospital episode statistics

## Declarations

### Ethical approval and consent to participate

This research has been conducted using the UK Biobank Resource under Application Number 82087, with ethical approval granted by the NHS North West Research Ethics Committee (REC reference 11/NW/0382). All participants gave informed consent to participate and be followed up through data-linkage.

### Consent for publication

Not applicable.

### Availability of data and materials

The datasets generated and/or analysed in this study will be made available for bona fide researchers who apply to use the UK Biobank dataset by registering and applying at http://www.ukbiobank.ac.uk/register-apply. R code used for data management and analysis are available here: https://github.com/alexgillett/MDD_T2D_HbA1c_LMM.

### Authors’ contributions

ACG, SPH, JT, CML and FC contributed to the conception of this study. ACG, SPH, JT and CML contributed to the study design. KGY and HG advised on using primary care records for the analysis of T2D patients. ACG, SPH and DH conducted the T2D validation, including PGS creation. ACG conducted the longitudinal analysis. ACG and SPH wrote the first draft, and CML and JT were major contributors to writing the manuscript. All authors reviewed the manuscript and approved the final version.

## Acknowledgements

This research has been conducted using the UKB Resource under Application Number 82087-we are grateful to all UKB staff and participants. The authors acknowledge use of the research computing facility at King’s College London, CREATE^43^. For the purpose of open access, the author has applied a Creative Commons Attribution (CC BY) licence to any Author Accepted Manuscript version arising.

