## Supplementary Materials for "The impact of major depressive disorder on glycaemic control in type 2 diabetes: a longitudinal cohort study using UK Biobank primary care records"

*Gillett et al.*

**Contents**

|  |  | Page |
| --- | --- | --- |
| 1. | Participant-level quality control for analysis with genetic data | 2 |
| 2. | Diabetic medicine extraction and variable creation | 2 |
| 3. | Type 2 diabetes diagnosis – exclusion criteria | 3 |
| 4. | Valid HbA1c | 3 |
| 5. | MDD diagnosis | 3 |
| 6. | BMI, SBP and DBP at T2D diagnosis | 3 |
| 7. | T2D polygenic score | 5 |
| 8. | Multiple imputation | 6 |
| 9. | Residual within-subject variability | 9 |
| 10. | Non-linear time-trend for post-T2D MDD individuals | 13 |

**Common accronyms used:**

T2D = type 2 diabetes

MDD = major depressive disorder

UKB = UK Biobank

BMI = body mass index

SBP = systolic blood pressure

DBP = diastolic blood pressure

MI = multiple imputation

GP = general practioner

SuppTable = Supplementary Table

SuppMethods = Supplementary Methods

SuppFigure= Supplementary Figure

**1. Participant-level quality control for analysis with genetic data**

When polygenic scores are utilised, this study had the following genetic quality control (QC) criteria for participant inclusion.

1) The individuals are of European ancestry based on:
   a. 4-means clustering on the first 2 principal components (provided by the UKB), and,
   b. 1000 Genomes Phase 3 projected principal components of population structure, keeping only those within three standard deviations from the mean for the first 100 principal components (as noted by Pain, et al (2021)^1^, this process removes individuals who are outliers due to technical genotyping or imputation errors).

Individuals are not closely related (>3^rd^ degree relatives were removed based on relatedness kinship estimates provided by the UKB)^2^.

**2. Diabetic medicine extraction and variable creation**

UK Biobank prescription data were coded using three different classification systems, Read v2, the British National Formulary (BNF) and the Dictionary of Medicines and Devices (dm+d). To extract relevant prescription records for glucose lowering medication, we used the Read v2 codes to extract prescriptions with a matching Read v2 code. BNF and dm+d data did have formatting differences within each classification system, and we therefore extract relevant prescription records based on drug name. SuppTable S3 contains the codes and drug names used to extract glucose lowering drugs.

Glucose lowering medication was used to help identify individuals with type 2 diabetes (T2D), as being prescribed a T2D medication forms part of the T2D diagnostic criteria used in this work. Although, being prescribed a glucose lowering medication makes an individual a possible T2D case, at least one other diagnostic criterion must be met (i.e. HbA1c measurement > 48 mmol/mol). Additonally, glucose lowering medication is part of the exclusion criteria too, whereby individuals prescribed more than one glucose lowering medication *at T2D diagnosis* are excluded from the analysis due to increased uncertainty over the validity of their extracted diagnosis date. See SuppMethods section 3 (next) for all exclusion criteria.

Diabetic medication is also used as a covariate in the longitudinal data analysis. Glucose lowering medication at each HbA1c observation was identified using prescription records up to three months prior to HbA1c measurement. This three-month boundary was used for two reasons:

1) HbA1c measures the average blood sugar levels over the previous 2-3 months. A prescription 3 months before an observation would therefore impact that HbA1c measurement.
2) The most frequent number of days’ supply prescribed in the UK for T2D medications is 28-days^3^. In most cases therefore our method will correctly identify the medication an individual is using at a given HbA1c observation.

**3. Type 2 diabetes diagnosis – exclusion criteria**

Individuals with a primary care code specific for type 1 diabetes were excluded. Individuals who had been treated with insulin in the first year after diagnosis or had a diagnosis before the age of 35 were excluded from the self-reported type 2 diabetes (T2D) diagnosis. Where clinical event or diabetic medication prescription date preceded or matched participant date of birth, it was in the year of their birth, or it was in the future, it has been altered by the UK Biobank (UKB) to predefined values in UKB data (01/01/1901, 02/02/1902, 03/03/1903 and 07/07/2037), and these values were set to missing for the analyses of this study^4^. People who either received insulin within the first year after their T2D diagnosis (*n*=246) or had more than 1 diabetic medication at the time of diagnosis (*n*=919) were removed from the sample. These people were either likely already receiving treatment prior to their recorded T2D diagnosis and it would be difficult to ascertain their actual date of diagnosis, or these individuals have a diagnosis of type 1 diabetes that has been coded incorrectly.

**4. Valid HbA1c**

HbA1c values in the primary care data consist of values based on the older DCCT method (based on % HbA1c) as well as values based on the newer IFCC method (mmol/mol). Values <3 and >197 were removed due to being outside the reportable range. For consistency, all values ≥3 and <15 were transformed to correspond with the IFCC method using the following formula^5^: IFCC HbA1c = (10.93 * DCCT HbA1c) - 23.5. There were 276 values between 15-19 (covering 210 individuals) for which it was not possible to determine the original unit of their HbA1c value and these values were therefore excluded from further analysis. To increase the number of HbA1c measurements available, HbA1c measurements from the UK Biobank biomarker data were combined with those from primary care. The biomarker HbA1c values were on average lower than those from primary care. To ensure consistency between we applied a correction to the biomarker HbA1c values^6^: calibrated HbA1c = (0.9696 * biomarker HbA1c) + 3.3595.

**5. MDD diagnosis**

UKB participants required at least two diagnostic codes for a depressive disorder or episode to be classed as MDD cases. Individuals with a diagnostic code for bipolar disorder, psychotic disorders or substance-use related disorders were excluded. Controls did not have any diagnostic code for depression, bipolar disorder, psychotic disorders or substance-use related disorders^7^.

**6. BMI, SBP and DBP at T2D diagnosis**

Extraction/ covariate details for all variables used/ explored in this analysis can be found in SuppTable 5a. How variables were used within the fixed effects model of the LDA is described in SuppTable 5b. This section describes: 1) data extraction for BMI, SBP and DBP and 2) creation of the variables- BMI, SBP and DBP at T2D diagnosis.

6.1. BMI at T2D diagnosis

*BMI extraction*

All available BMI observations, for all UK Biobank participants, were extracted from both primary care and UK Biobank assessments. These were then restricted to individuals diagnosed with T2D.

To extract BMI from primary care we used code ‘22K..’ to extract BMI measurements from read2 and read3 data. Additionally, we checked the following codes, Xa7wG and X76CO, however no BMI measurements were identified. Weight data from primary care records were also extracted using codes ‘22A..’, ‘1622’, ‘X76CG’, ‘XM01G’, ‘XE1h4’ and ‘Xa7wl’. Weights less than 30kg and greater than 200kg were removed^8^.

Extracting height from primary records was considered, however we were concerned about correctly identifying the correct unit of measurement for height records (CM, M, imperial measurements). Since we have an alternative available where the unit of measurement was known, namely height from the UK Biobank assessments, we decided to use this instead (data field 50). Additional BMI observations were then computed using GP record weight data and UK Biobank initial assessment height data.

BMI measurements taken at UK Biobank assessments were also extracted (data field 21001, all measurement instances).

All BMI measurements were combined into a dataset. We removed BMI observations less than 5 and greater than 200^9^. Duplicate entries were removed, as were those with event dates either in the future or at known incorrect dates for UK Biobank data: ‘1901-01-01’, ‘1902-02-02’, ‘1903-03-03’.

*Creating an approximate BMI at T2D diagnosis variable*

We added T2D diagnosis date for each individual into the BMI dataset. The time difference between each BMI observation and this diagnosis date was calculated. We restricted to BMI observations occurring up to 5 years before T2D diagnosis and 2 months after T2D diagnosis. We recognise that the included time frame prior to T2D diagnosis investigated is large at 5 years, however, we wished to retain as many people as possible in the analysis. Future work to develop methods including multiple time series in a multiple imputation (MI) algorithm (i.e. utilise BMI trajectories in addition to HbA1c trajectories in the MI algorithm) may overcome this limitation.

After restricting to observations within the window [-5 years, +2 months] around T2D diagnosis, we identified the observation closest to T2D diagnosis. If there were multiple observations identified for an individual we did the following:

1. Check if any of these observations occurred prior to T2D diagnosis. If yes, restrict to these. If not move to next step with all closest observations.
2. Then prioritise observation types in the following order: 1. UK Biobank records, 2. GP BMI records and 3. BMI calculated using GP weight data.
3. If there is still a tie retain the smallest BMI.

For individuals with an approximate BMI at T2D diagnosis available we then add this into the LDA dataset. Individuals where no BMI at T2D diagnosis is identified using the above approach are set as missing (NA) for this variable and imputed during the MI procedure for use in analysis 2 and 3. We note that BMI at initial UK Biobank assessment is included as an auxiliary variables in the MI model to improve imputation of BMI at T2D diagnosis.

6.2. SBP and DBP at T2D diagnosis

*Blood pressure extraction*

Blood pressure (BP) measures from GP records were extracted using code ‘246..’. BP measurements were extracted from UK Biobank assessment data using data fields 93 and 4080 for SBP and 94 and 4081 for DBP. For the GP records, if multiple BP measures were available on the same date, the maximum was used. Duplicate entries were removed, as were those with event dates either in the future or at known incorrect dates for UK Biobank data: ‘1901-01-01’, ‘1902-02-02’, ‘1903-03-03’. We also removed rows with dates ‘1958-12-15’ and ‘1960-01-01’ because these are considerably earlier than we would expect/ than other measurements available.

*Creating an approximate blood pressure at T2D diagnosis variables*

As with the extracted BMI data, we restricted to BP observations to those occurring up to 5 years before T2D diagnosis and 2 months after T2D diagnosis. We then identified the observation closest to T2D diagnosis. If there were multiple observations identified for an individual we took the mean of this tied data to be the BP measurement.

**7. T2D polygenic score**

T2D polygenic scores (PGSs) were created using T2D GWAS summary statistics from Scott et al (2017)^10^ (the discovery sample), and PRSice v2^11,12^. PRSice uses a p-value threshold and clumping PGS method. Clumping groups SNPs by linkage diseqillibruim (LD) blocks (here, clumping used r2 < 0.25 within a 250kb window) and selects the most significant SNP in that block (lowest discovery GWAS p-value). The resulting SNPs have their p-values compared to a p-value threshold. If the p-value is less than the threshold the SNP is included in the PGS. The PGS is then calculated in the target GWAS (here the eligible UKB sample of T2D cases and controls), where for each individual, their observed values for the selected SNPs are weighted by the correponding discovery GWAS SNP effect size, and then summed together. Multiple p-value thresholds are considered. For this study we used: 5.00E-08, 1.00E-05, 0.001, 0.01, 0.05, 0.1, 0.2, 0.3, 0.4, 0.5 and 1. Only genotyped SNPs with MAF > 0.01 were considered.

The 11 PGSs generated within the eligibile UKB sample by this process (one for each p-value threshold) were then used, in turn, in logistic regression with T2D case-control status as the outcome and the PGS as the regressor, adjusting for genetic batch, the first six principal components and assessment centre. More details for the general process used here can be found within Hagenaars, et al (2020)^13^.

The eligible UKB sample is outlined in the main text (with the individual-level genetic-based QC outlined in SuppMethods Section 1 above).

**8. Multiple imputation**

As described in the main text, HbA1c measurements occuring *after* T2D diagnosis are the outcome variables of interest in the longitudinal data analysis, with HbA1c measured *at* T2D diagnosis being used as a covariate in the longitudinal models for analysis 2 (adjusted mixed effects model) and analysis 3 (analysis of residual HbA1c variability). This HbA1c at T2D diagnosis measurement is not available for all participants. However, all patients have measurements occurring within a 6-month window of this date (a decision made as part of quality control to ensure that all individuals had a T2D diagnosis informative HbA1c measurement available).

In addition to there being missing data at HbA1c at T2D diagnosis, the following variables, intended for use in LDA, also had missing data (see Table 1 and SuppTable S12):

- BMI at T2D diagnosis (5.03% missing),
- SBP at T2D diagnosis (4.32% missing),
- DBP at T2D diagnosis (4.32% missing),
- Self-reported ethnicity (Other category selected in 1.4% of LDA study population),
- Qualifications (1.92% missing),
- TDI (0.27% missing),
- Ever-smoked (0.56% missing), and,
- Never consumed alcohol (0.24% missing).

{Note, that the % above are for the final LDA sample (i.e. not T2D sample or pre-imputation sample, see Figure 1 for distinctions).}

In order to include as many individuals as possible, we used the Amelia R package^14^ to impute missing data, with HbA1c at T2D diagnosis treated as a time series variable.

Amelia is a package that performs multiple imputation (MI) for time series data using a bootstrap based expectation-maximisation (EM) algorithm. It assumes that data is missing at random. Missing time series data can be predicted using polynomials of time or splines which allows non-linear time trends. Additionally, Amelia allows lags (previous measurement) and leads (next measurement) to be used to predict missing time series values.

The following variables were included in the MI model:

(1) HbA1c (mmol/mol) across time, including HbA1c measurements occurring **before, at** and **after** T2D diagnosis. Lags and leads for HbA1c were included. Note: for LDA, only measurements at and after T2D diagnosis are included.
(2) Time (relative to T2D diagnosis) is time since T2D diagnosis in years (T2D disease duration). HbA1c measurements occurring prior to T2D diagnosis will have a negative time value. Time splines with 3 knots were included.
(3) MDD exposure variables: MDD_index (pre-T2D MDD binary variable), MDD_change (time-varying post-T2D MDD binary variable), time between pre-T2D MDD and T2D diagnoses and time since post-T2D MDD diagnosis.
(4) All covariates to be included in analysis 2 and 3, such as BMI at T2D diagnosis, SBP at T2D diagnosis and DBP at T2D diagnosis (see SuppTable S5b).
(5) Auxiliary variables to improve imputation performance. These were:
    a. The T2D diagnostic criteria that an individual met (five binary variables for: HbA1c > 48 mmol/mol, T2D diagnostic code in GP data, T2D diagnostic code in HES data, prescribed T2D medication and self-reported T2D in UK Biobank).
    b. Genetically inferred ancestry (European, South Asian, East Asian, African, or Admixed African American- these are the 5 genetic super populations identified in the 1000 Genome project).
    c. Number of HbA1c observations occurring during study follow-up (10 years after T2D diagnosis).
    d. Number of HbA1c observations occurring prior to T2D diagnosis.
    e. Maximum SBP across all observations.
    f. Mean DBP at UK Biobank assessments.
    g. Number of BP measurements before T2D diagnosis.
    h. Maximum follow-up time for each participant.
    i. Additional pre-T2D MDD subgroup binary identifiers. At the point of MI we were unsure if we would consider a continuous time between pre-T2D MDD and T2D diagnosis (pre-T2D MDD duration) variable or if subgrouping using this time variable would be preferable. To keep our options open we decided to include a binary indicator for: 1) pre-T2D MDD duration <= 10^th^ percentile, 2) 25^th^ percentile < pre-T2D MDD duration <= 75^th^ percentile, and 3) pre-T2D MDD duration > 75^th^ percentile. The binary indicator variable for 10^th^ percentile < pre-T2D MDD duration <= 25^th^ percentile was excluded due to multicollinearity since we need to include a binary variable for pre-T2D MDD status at baseline.

Participant ID is used as the grouping variable.

The Amelia package typically assumes a multivariate normal distribution for the continuous variables considered in the model. This, of course, does not hold true for HbA1c at T2D diagnosis- patients diagnosed with T2D are typically above a HbA1c threshold (see SuppFigure S1; only ~3% of individiuals are observed to have HbA1c < 48 mmol/mol at diagnosis). Initially, we explored imputation models for HbA1c at T2D diagnosis by generating 10 test MI datasets for each of the following asumptions:

- HbA1c follows a normal distribution (no bounds).
- HbA1c follows a doubly truncated normal distribution with a lower truncation bound of 43 mmol/mol (with 43 being the 1^st^ percentile of the observed HbA1c at T2D diagnosis distribution) and an upper bound of 184 mmol/mol (184 being the maximum value observed for HbA1c at T2D diagnosis in the data available).
- HbA1c follows a doubly truncated log-normal distribution with lower truncation bound of log(43) and an upper bound of log(184).

A doubly truncated (log) normal distribution is not necessarily the most appropriate fit for the distribution of HbA1c measured at time points other than T2D diagnosis. However, for the HbA1c variable, we are only imputing HbA1c at diagnosis and therefore focus on generating imputed values that best match the distribution of measurements observed at this time point.

Supplementary Figure 9 presents a comparison of densities for: (a) the mean imputed values for HbA1c at T2D diagnosis from the model assuming HbA1c follows a normal distribution (red), and, (b) the observed HbA1c measurements at T2D diagnosis (turquoise). This plot shows that imputed HbA1c values at T2D diagnosis are, on average, lower than those observed, and the spread of the imputed distribution is wider around the mean.

Supplementary Figure S10 presents similar comparisons but for the imputation model assuming HbA1c at T2D diagnosis follows a doubly truncated normal distribution (lower truncation point at 43 mmol/mol and upper at 184 mmol/mol). Supplementary Figure S11 presents the same but for the doubly truncated log-normal distribution. Both improve on the imputation model assuming a normal distribution for HbA1c at T2D diagnosis. The doubly truncated normal distribution has a central tendency slightly greater for imputed compared to observed values. Whilst the doubly truncated log normal distribution has a central tendency slightly lower for imputed compared to observed values. We therefore decided to test one more lower truncation bound. This was 44.5 mmol/mol (~1.3 percentile of imputation population at T2D diagnosis)- selected with the aim of improving the log-normal distribution model by shifting the central tendency higher with a higher lower bound.

Supplementary Figure S12 plots the density of HbA1c at T2D diagnosis for the mean imputed values (red) and the observed measurements under the updated doubly truncated normal distribution assumption (i.e. lower truncation at 44.5 mmol/mol and upper at 184 mmol/mol). As expected, this model yields greater disparity between the observed and imputed densities- with the central tendency for the imputed data being higher than the observed. For the doubly truncated normal distribution, the lower bound set at 43 is preferred.

Supplementary Figure S13 plots the density of HbA1c at T2D diagnosis for the mean imputed values (red) and the observed measurements under the updated doubly truncated log-normal distribution assumption (i.e. lower truncation at log(44.5) mmol/mol and upper at log(184) mmol/mol). Whilst differences between the imputed and observed densities remain the tails of these distributions, the central tendencies are very similar, and this is the model, out those considered, that yields the best correspondence between imputed and observed densities for HbA1c at T2D diagnosis.

The imputation model selected to generate 50 MI datasets for use in analysis 2 and 3 was therefore the doubly truncated log-normal distribution with the lower truncation point set at log(44.5) and the upper at log(184). This is because, in the tests MI datasets generated, this model generated imputed HbA1c at T2D diagnosis that, on average, had the most similar distribution to the observed values.

Although selection of truncation points is arbitrary, we felt that:

- only 1.3% of observed dataset where less than the truncation point value,
- HbA1c at T2D diagnosis does have a lower bound for most people (the majority of people in the dataset will have HbA1c at T2D > 48), and,
- choosing the maximum observed value as the upper bound would avoid imputed predictions that were too high to be included in the analysis.

Although a log-normal model was used for imputation, HbA1c was transformed back to the original scale by Anmelia and this untransformed HbA1c used in the main analysis as we wanted model parameters to be on the untransformed scale for ease of interpretation.

Supplementary Figures S14 – S18 plot the densities for mean inputed values and observed values from the final imputation model for the following variables: HbA1c at T2D diagnosis, BMI at T2D diagnosis (standardised), TDI (standardised), SBP at T2D diagnosis (standardised) and DBP at T2D diagnosis (standardised). Note, for those with fraction missing presented, this fraction correspond to the pre-imputation sample.

**9. Residual within-subject variability**

The linear mixed effects models used for each individual ($i=1, \ldots, n$), are of the form:

$$\underline{Y}_{i}=\underline{X}_{i}\underline{\beta} +\underline{Z}_{i}\underline{b}_{i}+\underline{e}_{i}$$

Where, for individual *i*:

(a) $\underline{Y}_{i}$ is the vector of containing $T_{i}$ repeated HbA1c measurements.
(b) $\underline{e}_{i}$ is a vector of random error variables (random residuals) where $\underline{e}_{i} \sim N(\underline{0}, \underline{R}_{i})$, where $\underline{R}$ is the $T_{i}$ x $T_{i}$ covariance matrix.
(c) The fixed effects model is ‘$\underline{X}_{i}\underline{\beta}$’ where $\underline{X}_{i}$ is the fixed effects design matrix (containing the exposures, time splines, the interaction between exposure and the time splines and the additional covariates adjusted for- see SuppTables S5a and S5b), and $\underline{\beta}$ is the vector of fixed population parameters (fixed effects).
(d) $The$random effects model ‘$\underline{Z}_{i}\underline{b}_{i}$’ where $\underline{Z}_{i}$ is the random effects design matrix (containing random intercept and slope for time), and $\underline{b}_{i}$ is the vector of the 2 random effects, where it is assumed that $\underline{b}_{i} \sim N(\underline{0}, \underline{D})$ with $\underline{D}= \left[ \begin{matrix} \sigma_{1}^{2} & \sigma_{12} \\ \sigma_{21} & \sigma_{2}^{2} \end{matrix} \right]$ .

Marginally, the outcome variables are assumed to be normally distributed with mean equal to $\underline{X}_{i}\underline{\beta}$ and covariance matrix equal to $\underline{R}_{i}+ \underline{Z}_{i}\underline{D} \underline{Z}_{i}^{T}$.

Given the fixed and random effects (commonly known as conditionally), the outcome variables are still assumed to be normally distributed but with mean equal to $\underline{X}_{i}\underline{\beta} +\underline{Z}_{i}\underline{b}_{i}$ and covariance matrix equal to $\underline{R}_{i}$.

The residual covariance matrix for HbA1c, $\underline{R}_{i}$, captures within-individual variation. Recall from the main paper, analysis 1 uses mixed effects models to investigate the relationship between MDD exposures and HbA1c over time in individuals with T2D without adjusted for further covariates. In these models, we assume:

$$\underline{R}_{i}= \sigma_{R}^{2}\underline{\Lambda}_{i}$$

Where:

(a) $\sigma_{R}^{2}$ is the residual variance for individual *i*, and,
(b) $\underline{\Lambda}_{i}$ is correlation matrix for the residuals defined in this study as a continuous-time autoregressive order 1 (CAR1) error structure, with diagonal elements equal to 1, and off diagonal elements of $\underline{\Lambda}_{i}$ equal to $\phi^{|t_{ij}-t_{ik}|}$ ($\left| t_{ij}-t_{ik} \right|$ is the absolute value of the time difference between the two observations and $0<\phi<1$ is a parameter to be estimated).

This set-up for the residual within-individual covariance matrix is also used in analysis 2, which takes the preferred model from analysis 1 and updates it to include additional covariates in the fixed effects model.

Analysis 3 builds on this further by allowing residual within-individual variability in HbA1c to be a function of the binary MDD exposures. That is, the fixed effects and random effects model from analysis 2 are used but the residual variability is update to include binary MDD exposures. Three models are considered here:

1 **Residual variance is allowed to differ depending on MDD_index (i.e. whether an individual has a history of MDD or not at T2D diagnosis).**

Here we assume:

$$\underline{R}_{i}= \sigma_{R}^{2}e^{\theta_{pre}{MDD\_index}_{i}}\underline{\Lambda}_{i}$$

Where:

(a) ${MDD\_index}_{i}$ is the random variable for MDD history at T2D diagnosis, taking values 0 if individual *i* has no record of a MDD diagnosis at or before their T2D diagnosis date, and 1 otherwise,
(b) $\sigma_{R}^{2}$ is the residual variance if ${MDD\_index}_{i}$ equals 0 (the individual has no history of MDD at T2D diagnosis/ is part of the reference group),
(c) $\sigma_{R}^{2}e^{\theta_{pre}}$ is the residual variance if ${MDD\_index}_{i}$ equals 1 (the individual has pre-T2D MDD), where model parameter $\theta_{pre}$ allows pre-T2D MDD individuals to have a different within-individual variation in HbA1c to all other participants.
2 **Residual variance is allowed to differ depending on MDD_change (i.e. post-T2D MDD individuals allowed to have different residual variation *after* MDD diagnosis compared to all others).**

Here we assume that *j^th^* row and *k^th^* column of the residual covariance matrix is:

$$\underline{R}_{i}[j,k]=\sigma_{R}^{2}\sqrt{e^{\theta_{post}{MDD\_change}_{it_{ij}}}}\sqrt{e^{\theta_{post}{MDD\_change}_{it_{ik}}}}\phi^{|t_{ij}- t_{ik}|}$$

Where:

(a) ${MDD\_change}_{it_{ij}}$ (${MDD\_change}_{it_{ik}})$ is the time-varying random variable representing if individual *i* has been *newly* diagnosed with MDD between the start of follow-up and $t_{ij}$ ($t_{ik})$ for *j*, *k* = 1, 2, … $T_{i}$. That is: ${MDD\_change}_{it_{ij}}=\left\{ \begin{matrix} 1 if {MDD\_index}_{i}=0 and MDD diagnosed between T2D disease duration (0, t_{ij}] years \\ 0 otherwise \end{matrix} \right\}$
(b) $\sigma_{R}^{2}$ is the residual variance for individual *i* at time point $t_{ij}$ if ${MDD\_change}_{it_{ij}}=0$,
(c) $\sigma_{R}^{2}e^{\theta_{post}}$ is the residual variance for individual *i* at time point $t_{ij}$ if ${MDD\_change}_{it_{ij}}=1$, and,
(d) $\phi$ is the model parameter specifying the degree of autocorrelation between observations $t_{ij}$ and $t_{ik}$ (see CAR1 in analysis 1 description above).

$\theta_{post}$ is therefore the model parameter that allows individuals with post-T2D MDD to have a different within individual, residual HbA1c variance after their diagnosis with MDD compared to all others.

For this time-varying binary variable it is easier to specify $\underline{R}_{i}$ with a cell-specific specification as variance changes within an individual depending on if/ when they are diagnosed with MDD during study follow-up.

3 **Residual variance is allowed to differ depending on retrospective post-T2D MDD status (i.e. post-T2D MDD individuals allowed to have different residual variation compared to all others across *all* of follow-up).**

The residual variance specification in point 2 (directly above) allows individuals with post-T2D MDD to have different within-individual variation in HbA1c *after* their MDD diagnosis. We now define residual variation such that:

$$\underline{R}_{i}= \sigma_{R}^{2}e^{\theta_{1}E_{i}}\underline{\Lambda}_{i}$$

Where:

(a) $E_{i}$ is the binary random variable representing the post-T2D MDD group, taking value 0 if individual *i* is never diagnosed with MDD or is diagnosed at or prior to T2D diagnosis, and 1 if the individual is diagnosed with MDD after their T2D diagnosis.
(b) $\sigma_{R}^{2}$ is the residual variance if $E_{i}=0$ (that is pre-T2D MDD or no MDD/ the reference),
(c) $\sigma_{R}^{2}e^{\theta_{1}}$ is the residual variance if $E_{i}=1$, where model parameter $\theta_{1}$ allows residual variation to differ for the post-T2D MDD from the ‘no MDD’ and pre-T2D MDD groups across all follow-up, and,
(d) $\underline{\Lambda}_{i}$ is correlation matrix for the residuals defined in this study as a continuous-time autoregressive order 1 (CAR1) error structure, with diagonal elements equal to 1, and off diagonal elements of $\underline{\Lambda}_{i}$ equal to $\phi^{|t_{ij}-t_{ik}|}$ ($\left| t_{ij}-t_{ik} \right|$ is the absolute value of the time difference between the two observations and $0<\phi<1$ is a parameter to be estimated).

This set up for residual variation allows within-individual HbA1c variability to differ for post-T2D MDD individuals, compared to all others, across all of follow-up, potentially identifying if some differences start prior to MDD diagnosis.

Note, the above is all looking within an individual *i*, and we assume that there is no correlation between different individuals.

For more information please see Pinheiro and Bates (2000)^15^ and Pinheiro, Bates and R Core Team (2022)^16^. Harrell (2001)^17^ also provides a good overview of residual variance-covariance structures for longitudinal data analysis (focusing on generalised least squares, so no random effects considered).

If of interest, restricted cubic splines are also well explained in Harrell (2001)^17^, and Gauthier, et al (2020)^18^.

**10. Non-linear time-trend for post-T2D MDD individuals**

In the main manuscript we presented results from mixed effects models allowing the HbA1c-trend for post-T2D MDD individuals to differ from no MDD by a linear time-since-MDD diagnosis variable. In addition to this, although not presented in the main results primarily due to space restrictions, we also ran unadjusted models where time-since-MDD diagnosis for post-T2D MDD individuals was allowed to be non-linear, by considering a restricted cubic spline (RCS) with 4 knots for this variable. However, when comparing the model with this non-linear time-since-MDD effect to the model with the linear time-since-MDD effect using a likelihood ratio test, the linear model was preferred (*p* = 0.2111). Additionally, adding the extra post-T2D MDD duration parameters did not improve the AIC (the linear duration model had AIC = 247786.0 compared to AIC = 247786.8 for the non-linear model), and the linear model was preferred when looking at the BIC (BIC = 247915.0 for the linear post-T2D MDD duration model, BIC = 247935.7 for the non-linear post-T2D MDD duration model). However, the available sample size for this group n=226, and further investigation into the change in HbA1c trends after a MDD diagnosis for individuals with T2D using a larger sample size (i.e. using CPRD) may yet reveal a non-linear effect.
