## Supplementary Figures for "The impact of major depressive disorder on glycaemic control in type 2 diabetes: a longitudinal cohort study using UK Biobank primary care records"

Table of content

|  | Page |
| --- | --- |
| **Plots corresponding to Descriptive Analysis** | 2 |
| S1: Density plot of HbA1c (mmol/mol) at T2D diagnosis by MDD subgroup. | 2 |
| S2: Density plot of age (years) at T2D diagnosis by MDD subgroup. | 3 |
| S3: Density plots stratified by MDD subgroup for follow-up time (T2D disease duration in years). | 4 |
| S4: Density plots stratified by MDD subgroup for number of pre-T2D measurements. | 5 |
| S5: Density plot of approximate BMI at T2D diagnosis by MDD subgroup. | 6 |
| S6: Density plot of approximate SBP at T2D diagnosis by MDD subgroup. | 7 |
| S7: Density plot of approximate DBP at T2D diagnosis by MDD subgroup. | 8 |
| S8: Density plot of TDI by MDD subgroup. | 9 |
| **Plots corresponding to Multiple Imputation** | 10 |
| S9: Density plots comparing the distribution of HbA1c at T2D diagnosis for observed values versus imputed values when the imputation model assumes normality for HbA1c (10 test MI datasets) | 10 |
| S10: Density plots comparing the distribution of HbA1c at T2D diagnosis for observed values versus imputed values when the imputation model assumes a doubly truncated normal distribution for HbA1c (bounds = [43, 184] mmol/mol; 10 test MI datasets) | 11 |
| S11: Density plots comparing the distribution of HbA1c at T2D diagnosis for observed values versus imputed values when the imputation model assumes a doubly truncated log-normal distribution for HbA1c (bounds = [log(43), log(184)]; 10 test MI datasets) | 12 |
| S12: Density plots comparing the distribution of HbA1c at T2D diagnosis for observed values versus imputed values when the imputation model assumes a doubly truncated normal distribution for HbA1c (bounds = [44.5, 184] mmol/mol; 10 test MI datasets) | 13 |
| S13: Density plots comparing the distribution of HbA1c at T2D diagnosis for observed values versus imputed values when the imputation model assumes a doubly truncated log-normal distribution for HbA1c (bounds = [log(44.5), log(184)]; 10 test MI datasets) | 14 |
| S14: Density plots of observed values versus imputed values for HbA1c at T2D diagnosis, using final imputation model- doubly truncated log-normal distribution for HbA1c (bounds = [log(44.5), log(184)]; 50 MI datasets) | 15 |
| S15: Density plots of observed values versus imputed values for BMI at T2D diagnosis (standardised) | 16 |
| S16: Density plots of observed values versus imputed values for TDI (standardised) | 17 |
| S17: Density plots of observed values versus imputed values for SBP at T2D diagnosis (standardised) | 18 |
| S18: Density plots of observed values versus imputed values for DBP at T2D diagnosis (standardised) | 19 |

**Descriptive plots**

**Supplementary Figure S1:** Density plot of HbA1c (mmol/mol) at T2D diagnosis by MDD subgroup.

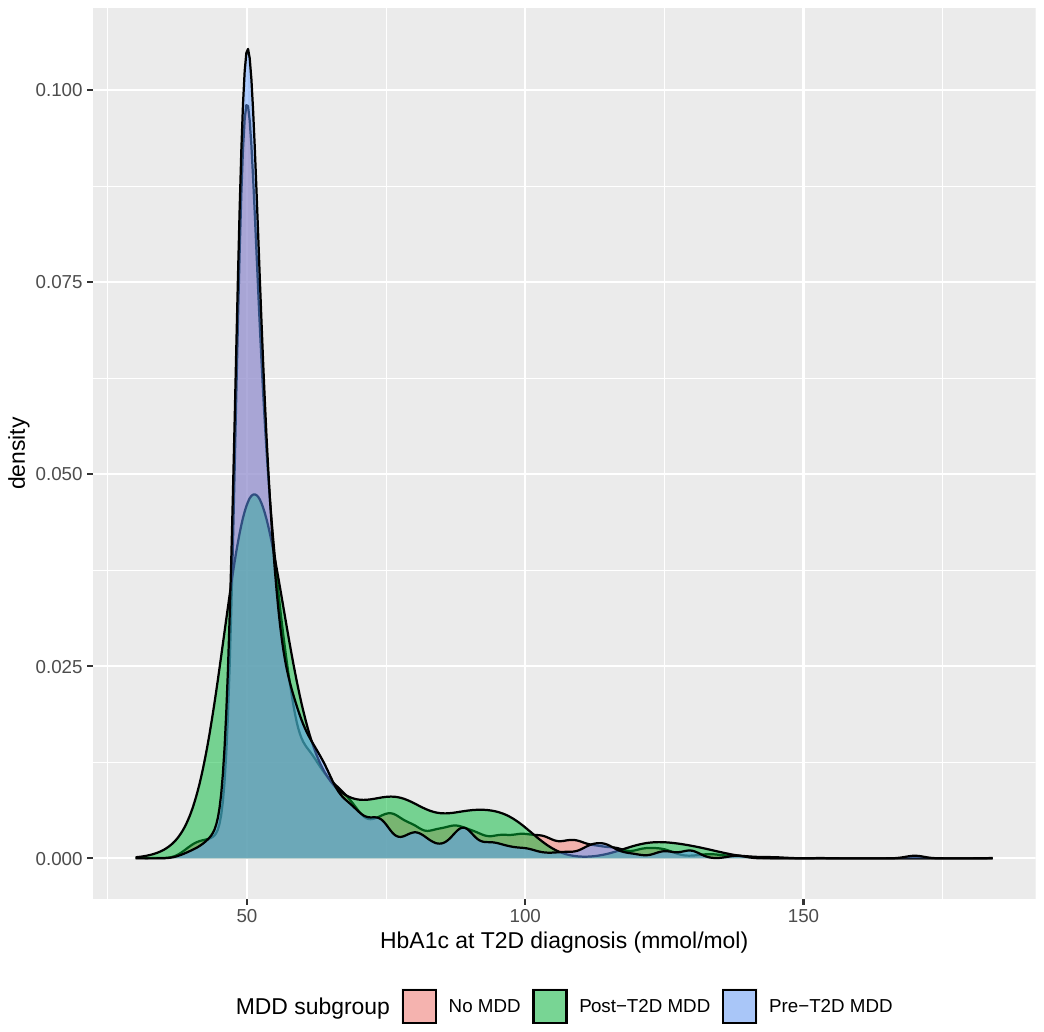

**Supplementary Figure S2:** Density plot of age (years) at T2D diagnosis by MDD subgroup.

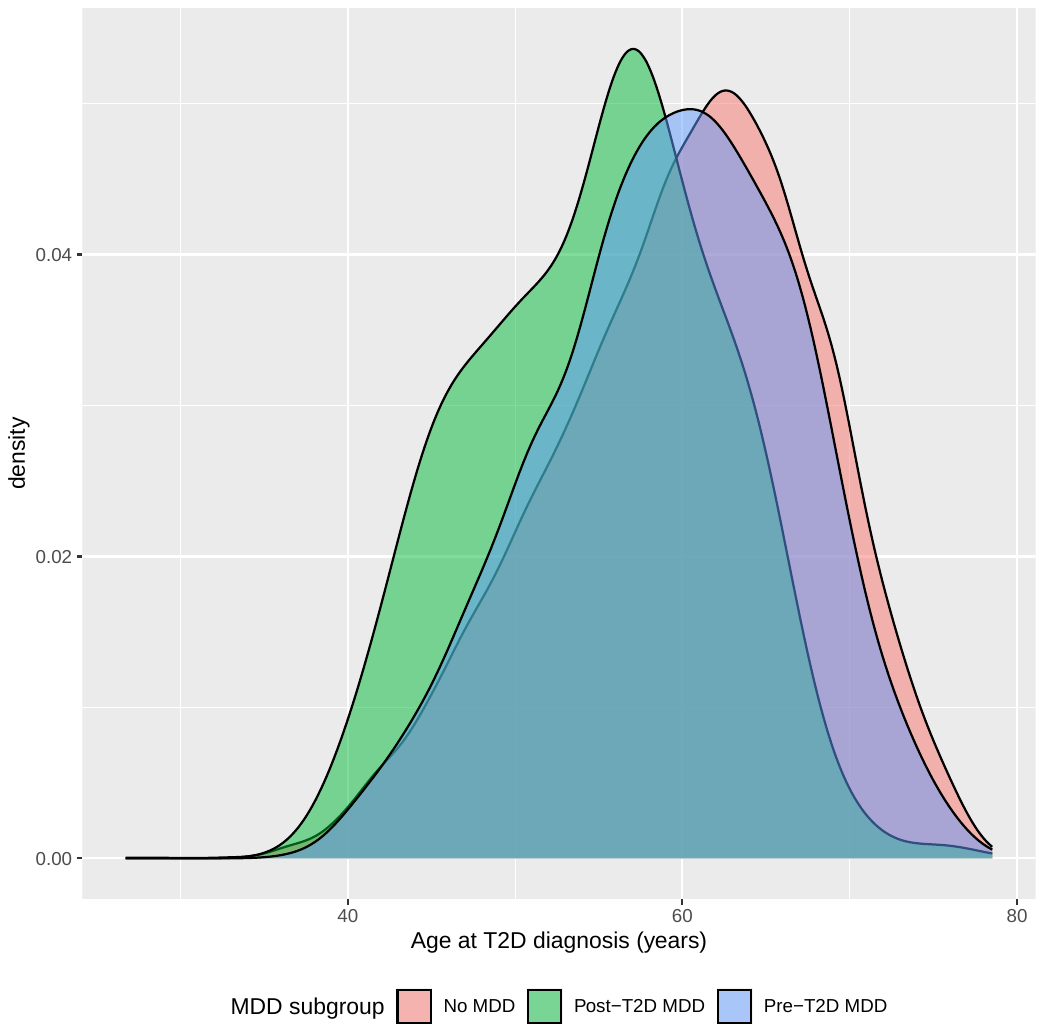

**Supplementary Figure S3:** Density plots stratified by MDD subgroup for follow-up time (T2D disease duration in years) up to 10 years after T2D diagnosis (end of study).

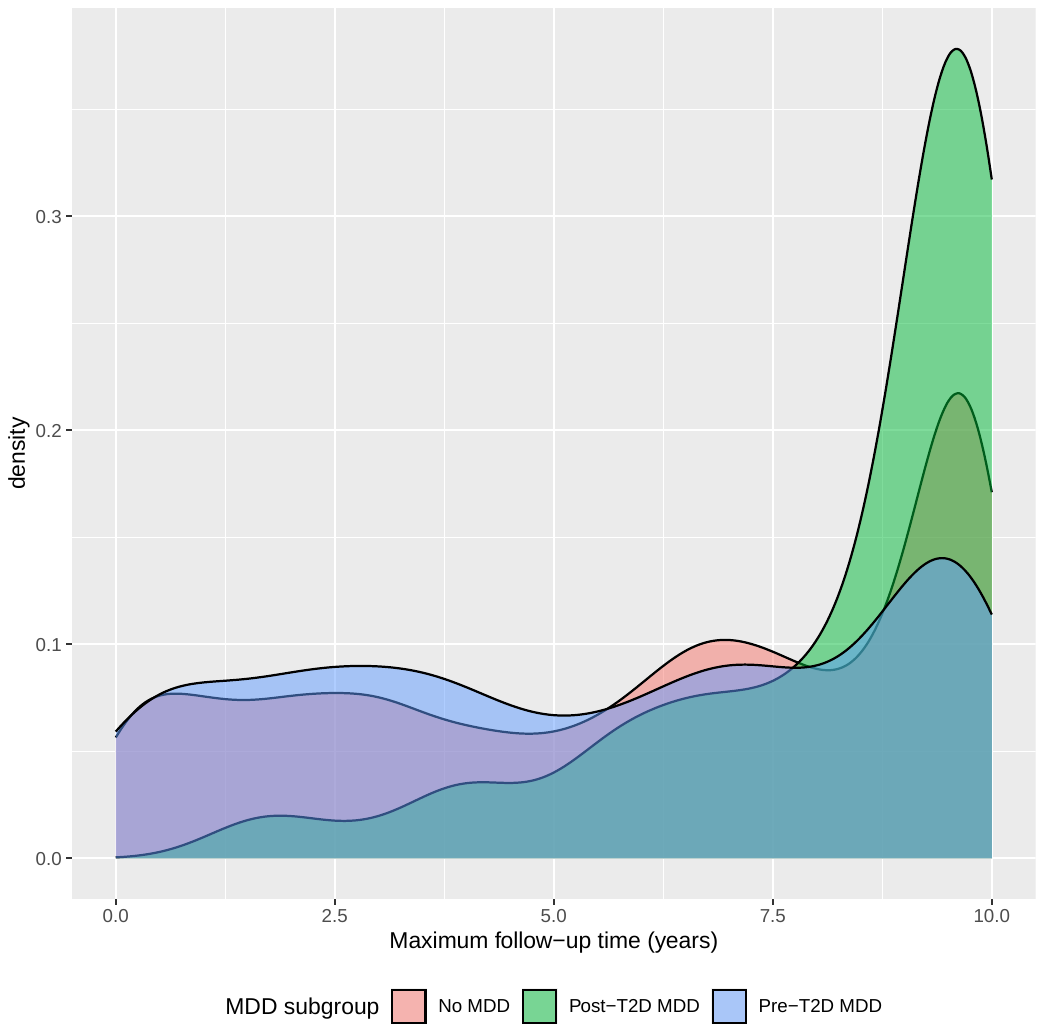

**Supplementary Figure S4:** Density plots stratified by MDD subgroup for total number of HbA1c, BMI and blood pressure measurements prior to T2D diagnosis.

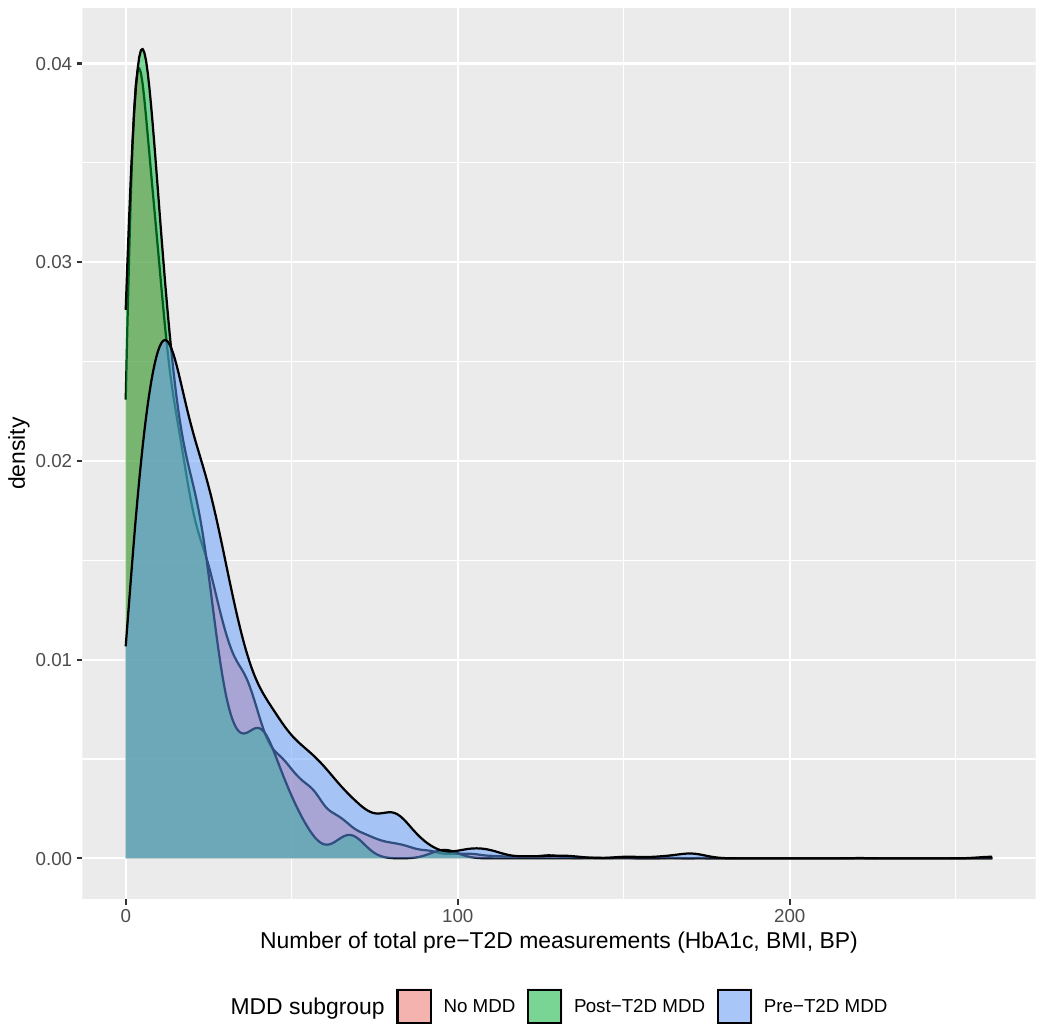

**Supplementary Figure S5:** Density plot of BMI (approximately) at T2D diagnosis by MDD subgroup.

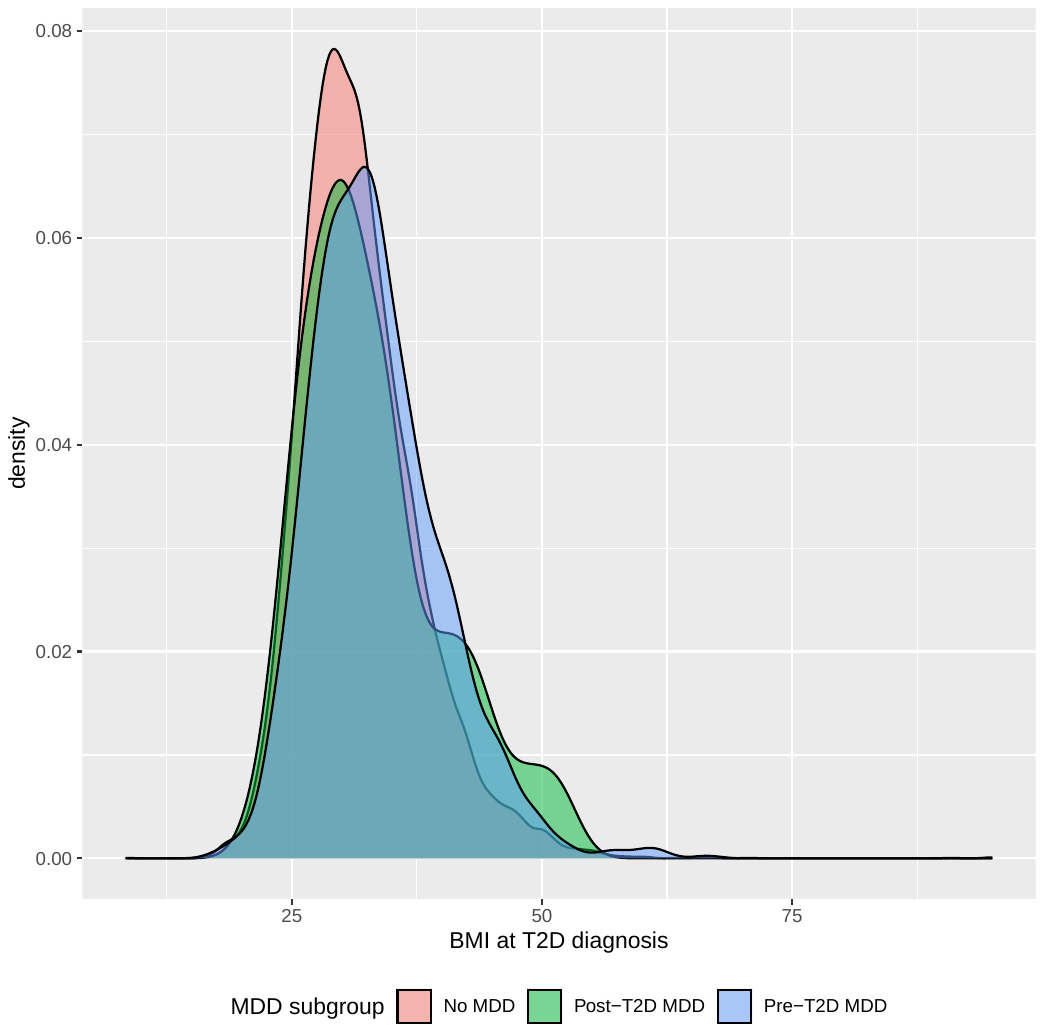

**Supplementary Figure S6:** Density plot of systolic blood pressure (approximately) at T2D diagnosis by MDD subgroup.

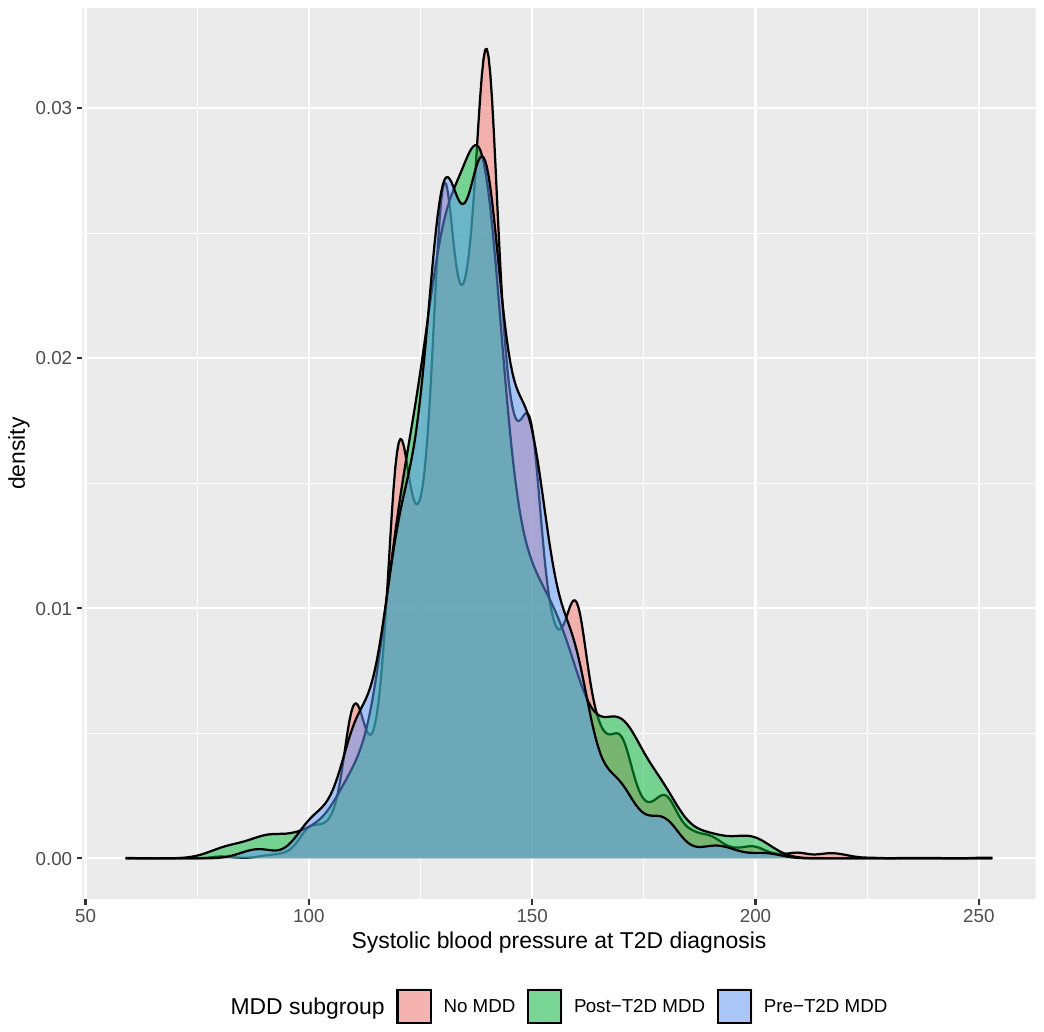

**Supplementary Figure S7:** Density plot of diastolic blood pressure (approximately) at T2D diagnosis by MDD subgroup.

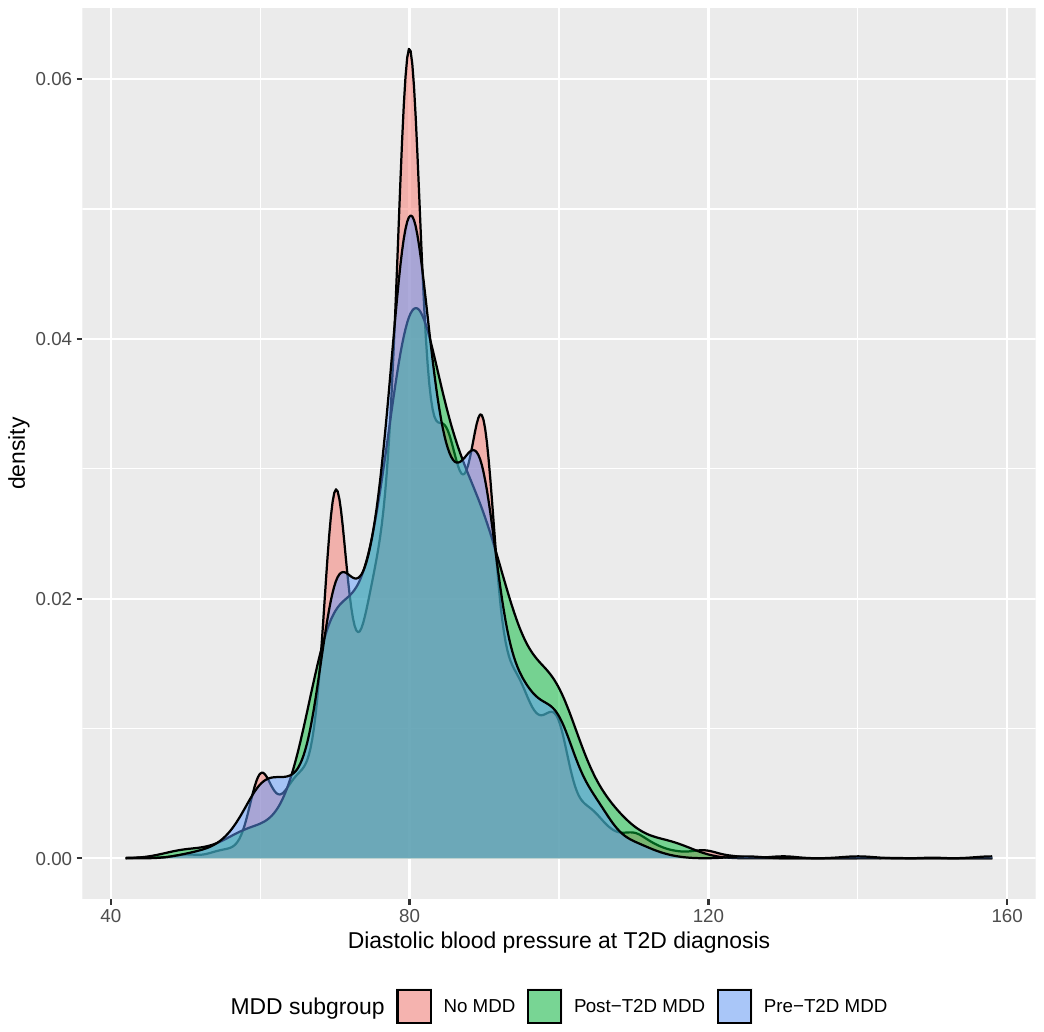

**Supplementary Figure S8:** Density plot of Townsend Deprivation Index (TDI) measured at initial UK Biobank assessment, by MDD subgroup.

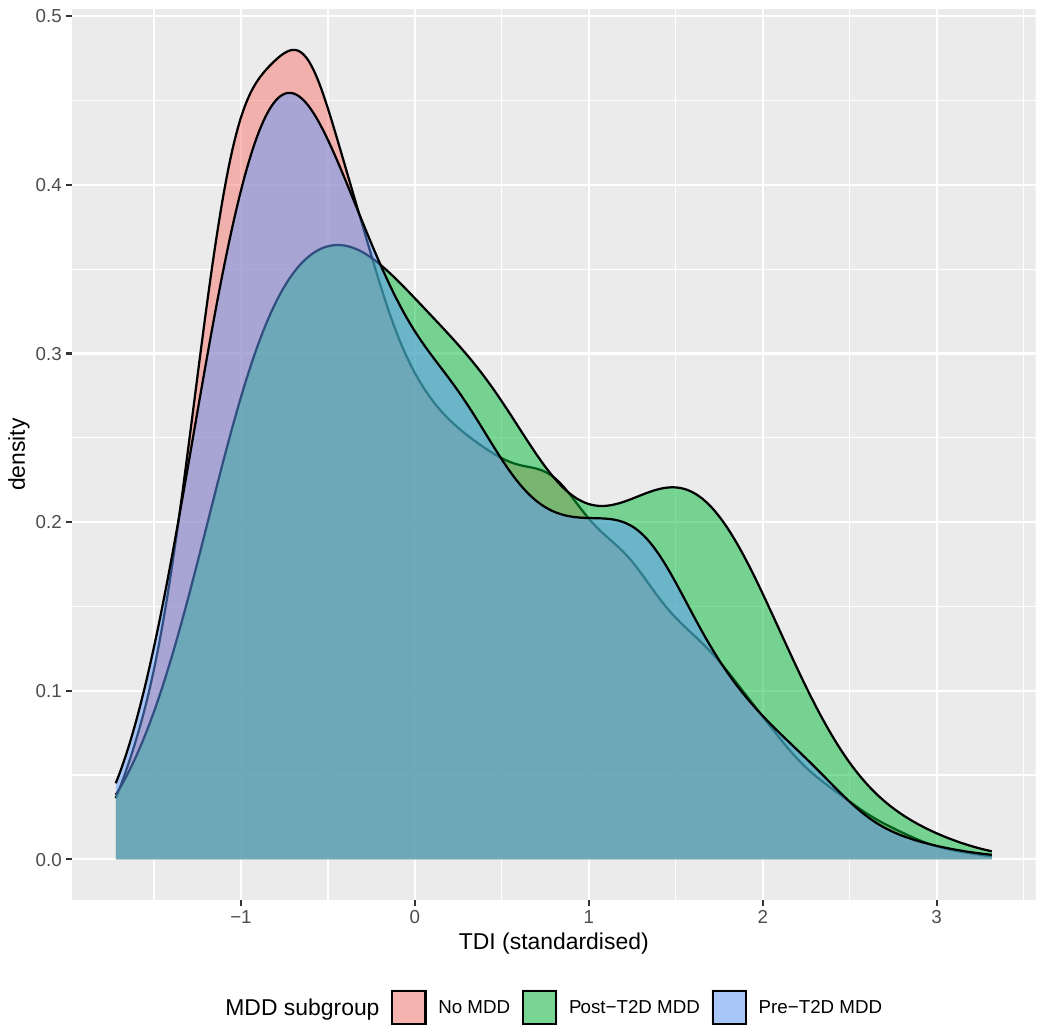

**Multiple imputation plots**

**Supplementary Figure S9:** Density plot for standardised HbA1c at T2D diagnosis for measured/ observed (turquoise) and imputed (red) values when the multiple imputation (MI) model assumes a **normal** distribution for HbA1c. Generated using 10 MI datasets. Imputation model tends to assign a lower value to imputed data compared to observed.

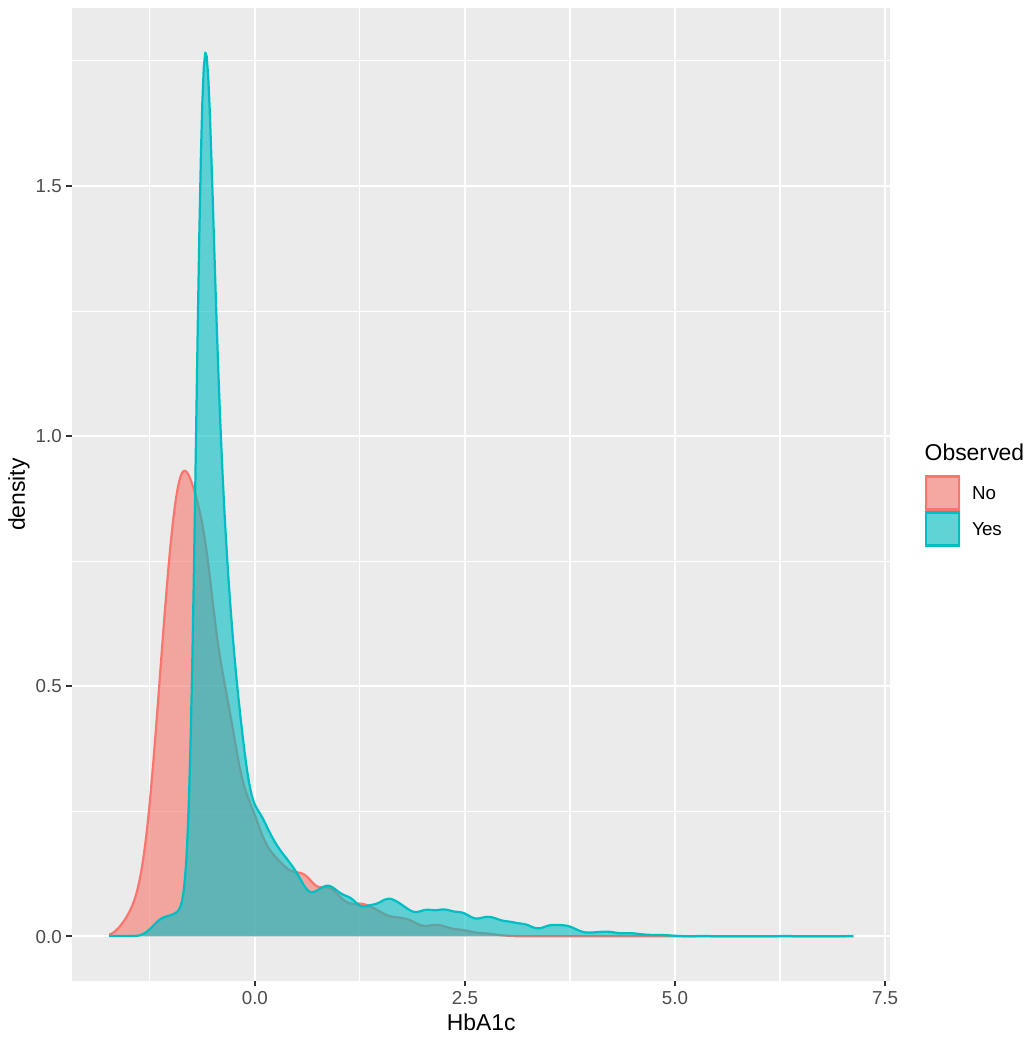

**Supplementary Figure S10:** Density plot for standardised HbA1c at T2D diagnosis for measured/ observed (turquoise) and imputed (red) values when the multiple imputation (MI) model assumes a **doubly truncated normal** distribution for HbA1c. Lower truncation is at 43 mmol/mol (-0.98 when standardised) and upper is at 184 mmol/mol (7.12 when standardised). Generated using 10 MI datasets.

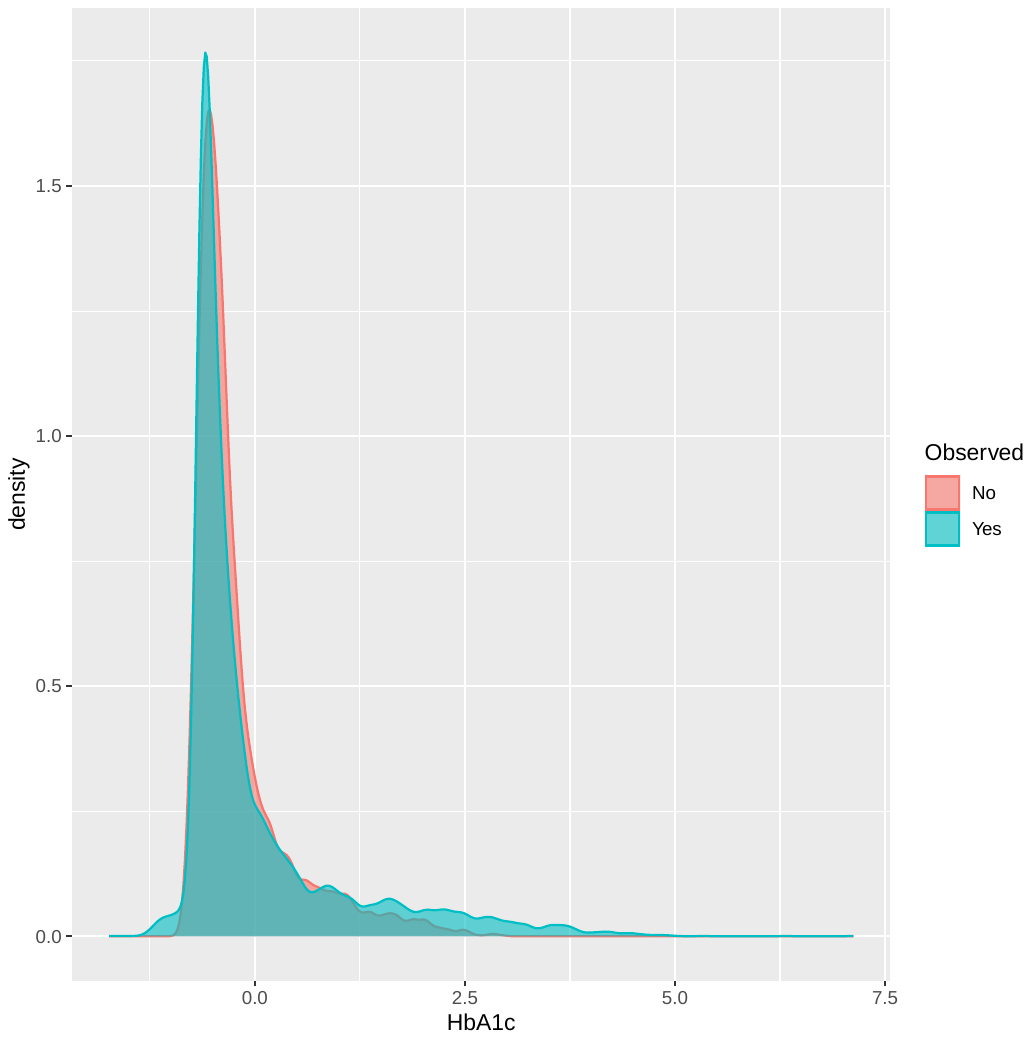

**Supplementary Figure S11:** Density plot for HbA1c at T2D diagnosis for measured/ observed (turquoise) and imputed (red) values when the multiple imputation (MI) model assumes a **doubly truncated** **log-normal** distribution for HbA1c. Lower truncation is at log(43) and upper is at log(184). Generated using 10 MI datasets.

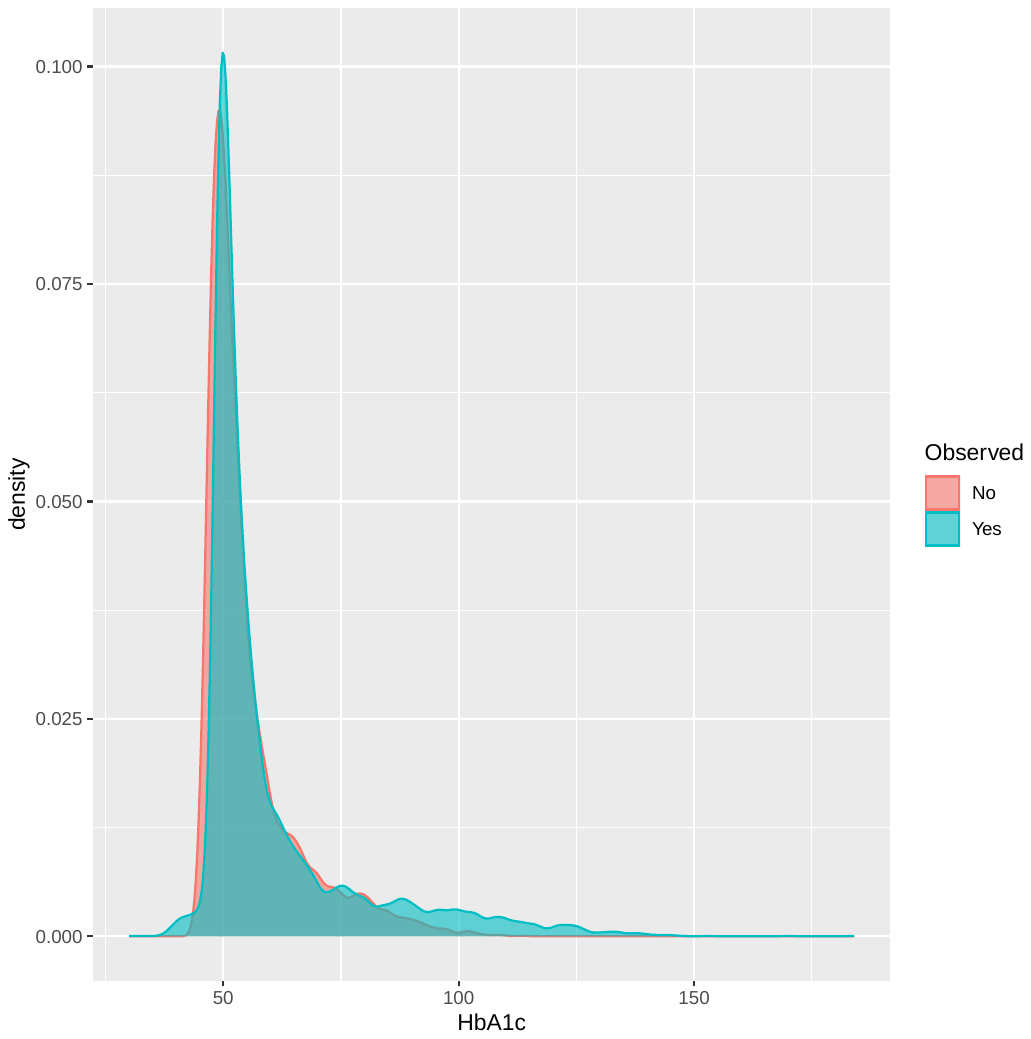

**Supplementary Figure S12:** Density plot for standardised HbA1c at T2D diagnosis for measured/ observed (turquoise) and imputed (red) values when the multiple imputation (MI) model assumes a **doubly truncated normal** distribution for HbA1c. Lower truncation is at 44.5 mmol/mol (-0.90 when standardised) and upper is at 184 mmol/mol (7.12 when standardised). Generated using 10 MI datasets.

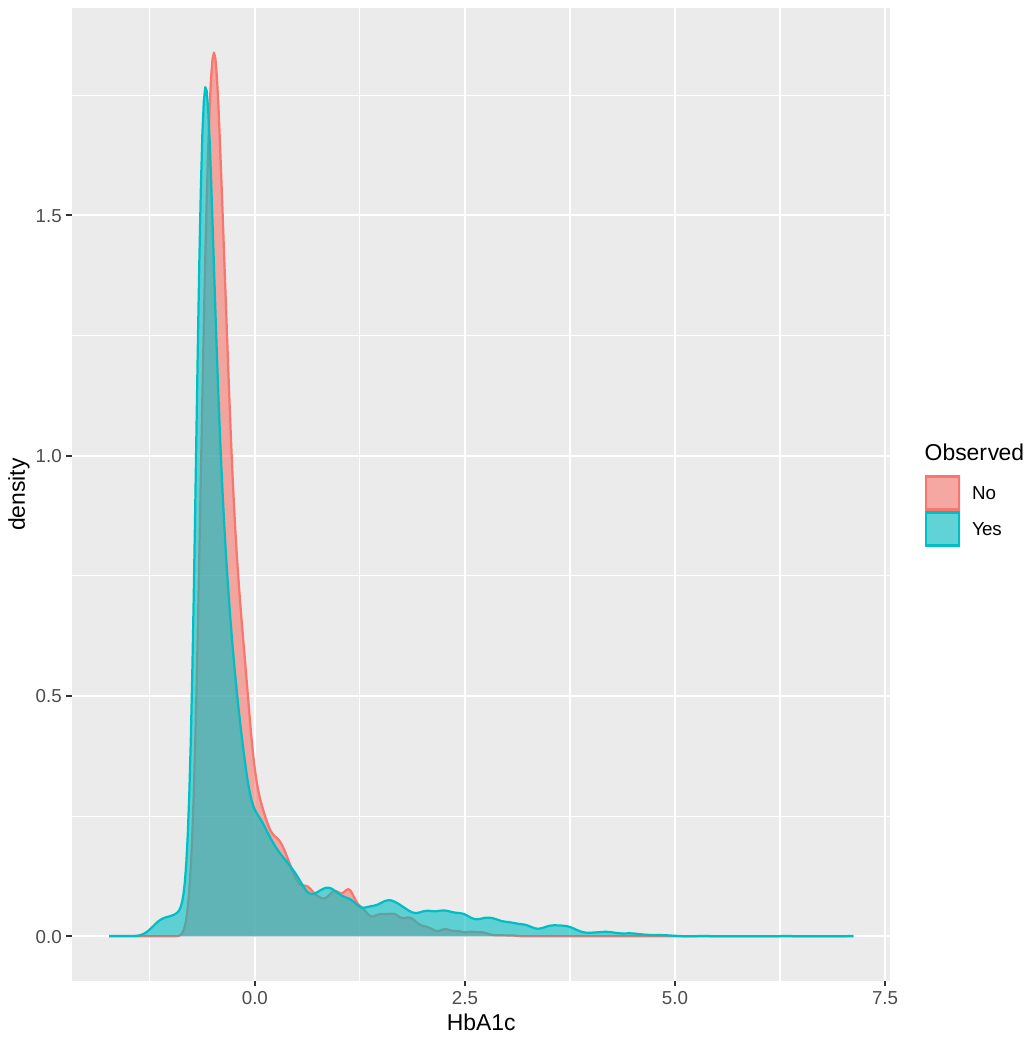

**Supplementary Figure S13:** Density plot for HbA1c at T2D diagnosis for measured/ observed (turquoise) and imputed (red) values when the multiple imputation (MI) model assumes a **doubly truncated** **log-normal** distribution for HbA1c. Lower truncation is at log(44.5) and upper is at log(184). Generated using 10 MI datasets.

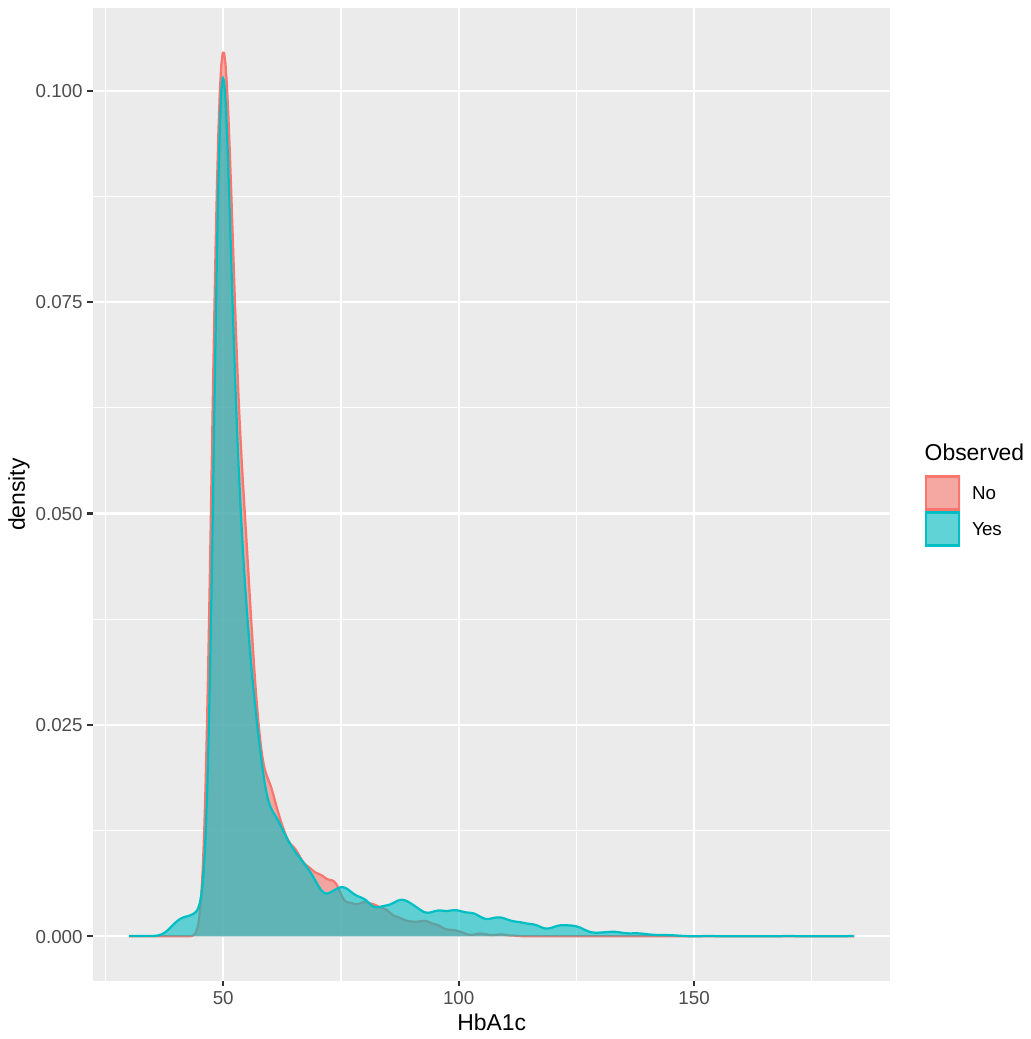

**Supplementary Figure S14:** Density plot for HbA1c (mmol/mol) at T2D diagnosis for measured/ observed (turquoise) and imputed (red) values when the multiple imputation (MI) model assumes a **doubly truncated** **log-normal** distribution for HbA1c. Lower truncation is at log(44.5) and upper is at log(184). Generated using 50 MI datasets. These are the MI datasets to be used in the main analysis.

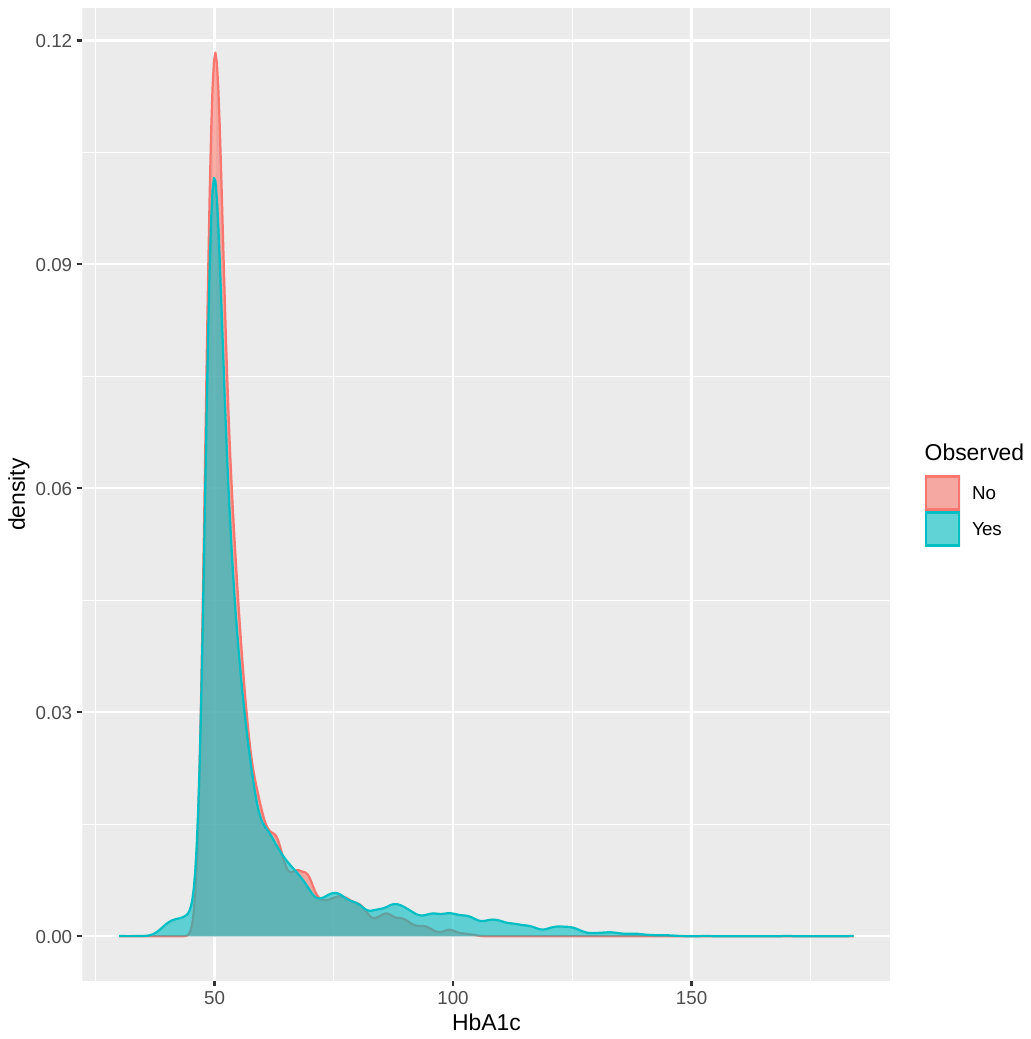

**Supplementary Figure S15:** Density plot for BMI at T2D diagnosis (standardised, bmi_std) for measured/ observed (blue) and imputed (red) values. Generated using 50 MI datasets. These are the MI datasets to be used in the main analysis.

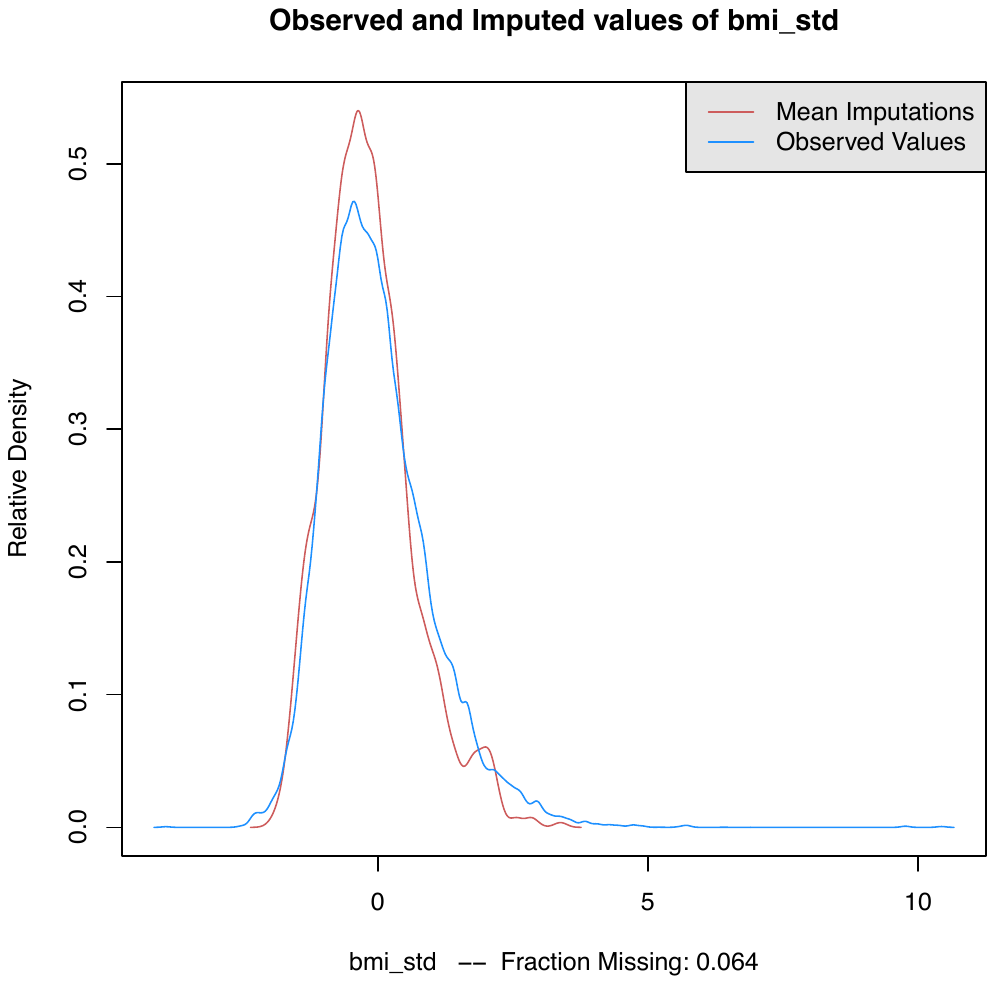

**Supplementary Figure S16:** Density plot for TDI (standardised, tdi_std) for measured/ observed (blue) and imputed (red) values. Generated using 50 MI datasets. These are the MI datasets to be used in the main analysis.

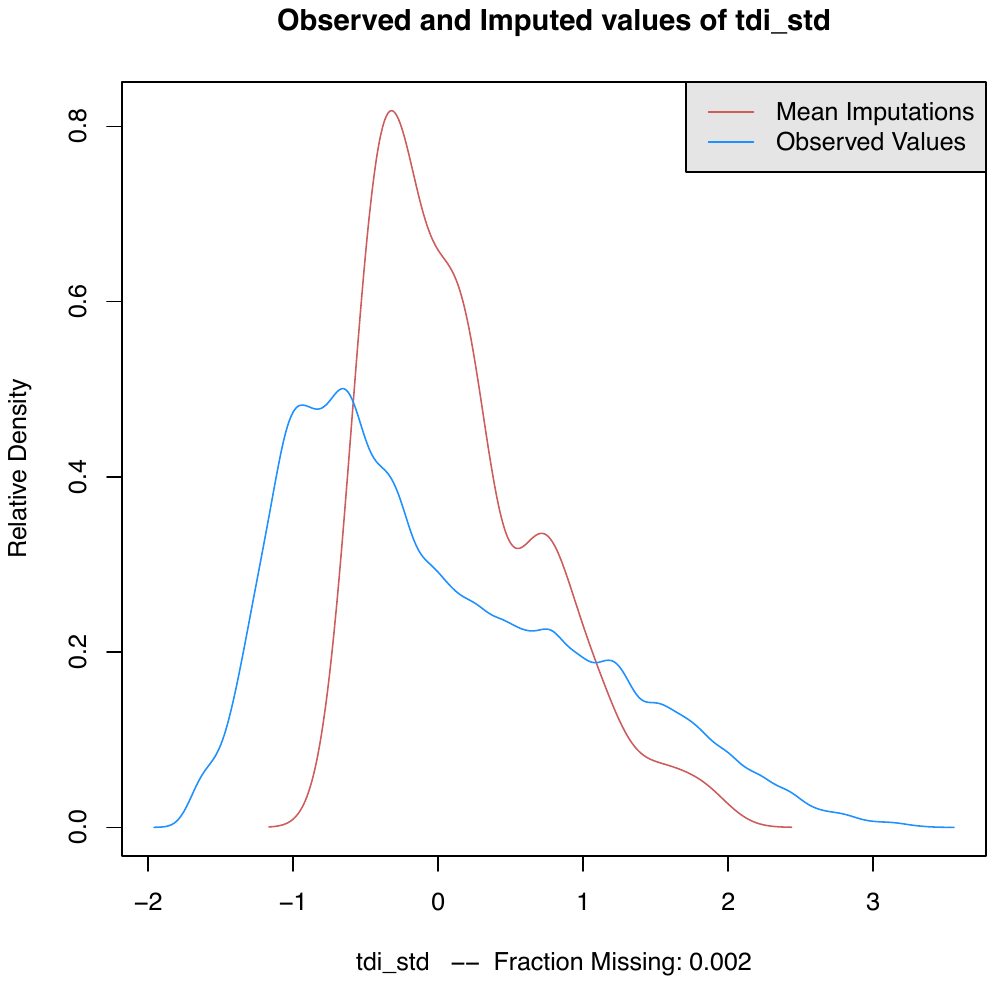

**Supplementary Figure S17:** Density plot for SBP at T2D diagnosis (standardised, sbp_base_std) for measured/ observed (blue) and imputed (red) values. Generated using 50 MI datasets. These are the MI datasets to be used in the main analysis.

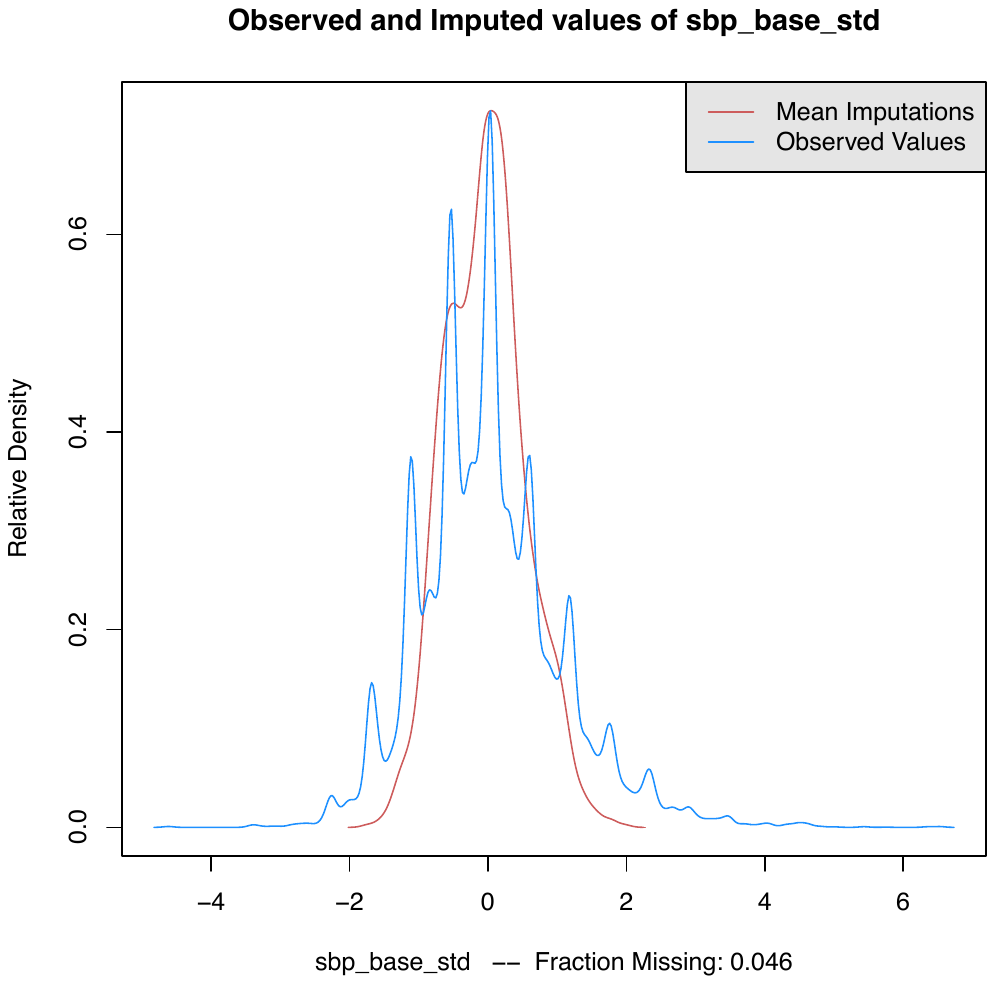

**Supplementary Figure S18:** Density plot for DBP at T2D diagnosis (standardised, dbp_base_std) for measured/ observed (blue) and imputed (red) values. Generated using 50 MI datasets. These are the MI datasets to be used in the main analysis.

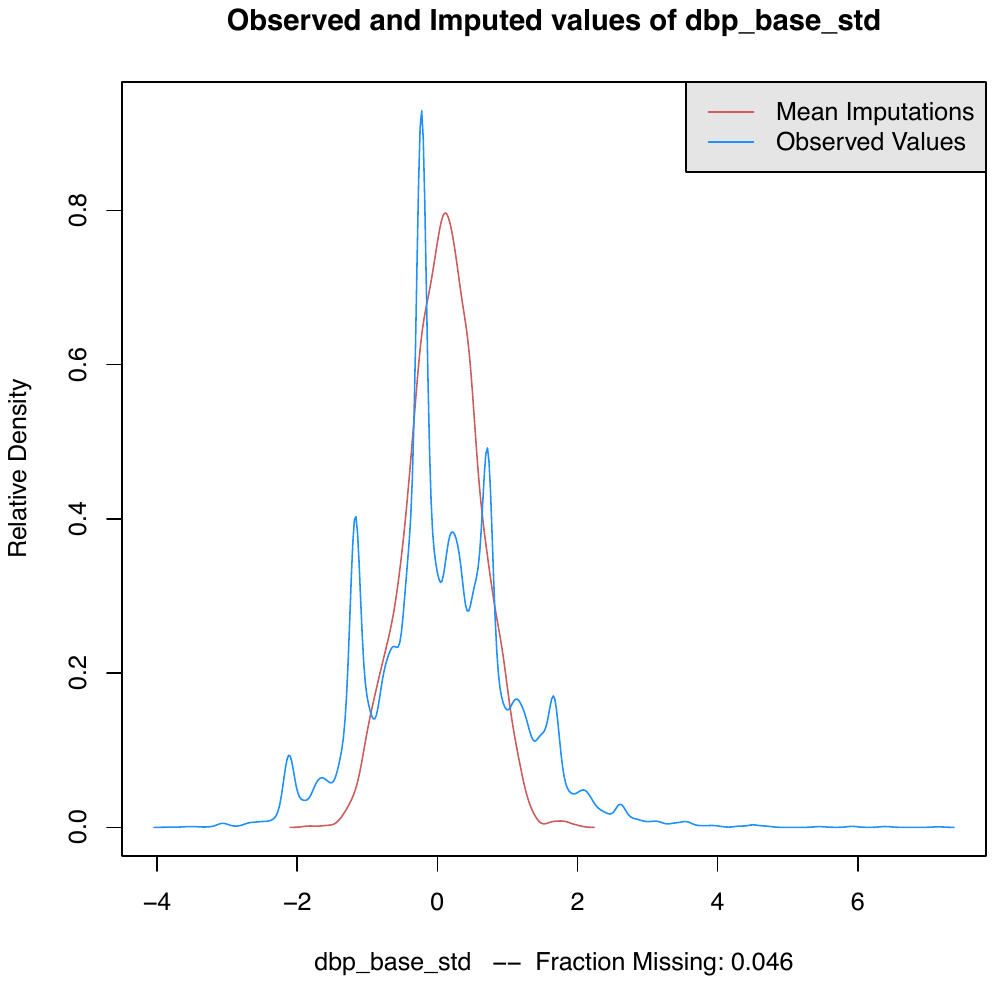
